# Exploring linkages between drought and HIV treatment adherence in Africa: A Systematic Review

**DOI:** 10.1101/2020.09.03.20187591

**Authors:** Kingsley Orievulu, Sonja Ayeb-Karlsson, Sthembile Ngema, Kathy Baisley, Frank Tanser, Nothando Ngwenya, Janet Seeley, Willem Hanekom, Kobus Herbst, Dominic Kniveton, Collins Iwuji

## Abstract

Climate change is directly and indirectly linked to human health, including through access to treatment and care. Our systematic review presents a ‘systems’ understanding of the nexus between drought and antiretroviral treatment (ART) adherence in HIV-positive individuals in the African setting. Narrative synthesis of 111 studies retrieved from Web of Science, PubMed/Medline, and PsycINFO suggests that economic and livelihoods conditions, comorbidities and ART regimens, human mobility, and psycho-behavioural dispositions and support systems interact in complex ways in the drought-ART adherence nexus in Africa. Economic and livelihood-related challenges appear to impose the strongest impact on human interactions, actions and systems that culminate in non-adherence. Indeed, the complex pathways identified by our systems approach emphasise the need for more integrated research approaches to understanding this phenomenon and develop interventions.

## 1. Introduction

A recent World Health Organisation (WHO) report supporting the negotiations of the United Nations Framework Convention on Climate Change (UNFCCC) recognised that climate change affects human health both directly and indirectly.^1^ The report notes an estimated close to 13 million deaths – about 23% of all global deaths – linked to modifiable environmental factors, often related to climate change.^2^ While direct health impacts of climate change, including physiological effects of exposure to higher temperatures and increasing incidence of Non-Communicable Diseases (NCDs), such as respiratory and cardiovascular diseases, are relatively well-understood,^3–6^ indirect effects on health, particularly those resulting from long causal pathways, such as through impacts on livelihoods, are more difficult to identify.

Drought is a major consequence of anthropogenic climate change, with impacts on human health. An already common phenomenon in Southern Africa, its frequency and duration are likely to increase with impact on health exacerbated by the region’s low adaptive capacity.^7–9^ Indeed, social vulnerabilities, especially high HIV prevalence and unemployment levels, further complicate the ways in which drought, and other climate and weather conditions, affect populations in this region.^9,10^

HIV prevalence in Southern Africa accounts for more than 30% of the global number.^11^ Both direct factors (sexual behaviour and unprotected sex) and longer causal chains (lack of adherence to HIV antiretroviral treatment (ART)) contribute to this high burden.^7,8^ Poor ART adherence increases morbidity and mortality risks in HIV-positive individuals as well as HIV transmission.^7,8^ We here focus on the indirect impacts of drought on HIV treatment adherence in Africa, with implications for Southern Africa.

The objective of this paper is therefore to develop a “systems” understanding of the nexus between environmental stress, in this case drought, and ART adherence in HIV-positive individuals in Africa.

The previously shown correlation between drought and increased HIV prevalence could be explained by changes in behaviour in reaction to income and production shocks, which often culminate in increased sexual risk taking, temporary migration, school withdrawal and early sexual debut, especially in rural contexts.^7,8^ However, to date no study has applied a full systems approach to understand this association, in particular to account for the complexities underlying drought’s impact on HIV ART adherence. Berry et al defines systems thinking as a set of ‘synergistic analytic skills’ used to help describe a complex set of interacting factors that produce outcomes, to predict their behaviour and to formulate interventions to achieve desired results.^12^ We argue that the systems approach is more appropriate in investigating the drought-HIV nexus as it shows how different geopolitical, socioeconomic and environmental factors, including health systems interact through a complex interlinked process and culminate in non-linear outcomes such as mental (ill)-health, or in our case HIV (non)-adherence.^12–14^ Thus, we undertook a systematic literature review on the impacts of drought on human health and livelihoods, and factors associated with ART adherence, and used the findings to develop a systems diagram describing the relationship between drought and ART adherence. This makes a case for future research agendas and policy frameworks that capture the complex causal pathways between drought and ART adherence especially in Africa.

## 2. Methods

Four researchers (KO, SAK, DK and CI) searched three electronic databases for peer-reviewed published literature: Web of Science, PUBMED/MEDLINE and PsycINFO (1 January 2003 – 20 September 2019). We chose 2003 as our starting date as it was about the time ART roll-out was starting in Africa.^15^ Three distinct searches were applied to each of the databases in line with the study objective to cover publications on impacts of drought generally, on human health and adherence to ART, in the African setting (Table 1A & Supp. Table 1). We reviewed primary studies published in English.

**Table 1.** Search strategy, inclusion and exclusion criteria.

| **A: Search Criteria** |  |  |
| --- | --- | --- |
| **Impact of drought** | **Drought and health** | **ART adherence** |
| Drought | Drought | HIV or Human Immunodeficiency Virus or AIDS or Acquired Immuno-Deficiency Syndrome |
| AND | AND | AND |
| Impact | Health | antiretroviral or ART or ARV |
| AND | AND | AND |
| Africa | Africa | viral load or RNA |
|  |  | AND |
|  |  | adherence or compliance or non-adherence or noncompliance or treatment adherence or treatment compliance |
|  |  | AND |
|  |  | Africa |
| **B. Inclusion and Exclusion Criteria** |  |  |
| **Inclusion criteria** |  | **Exclusion criteria** |
| HIV+ |  | HIV- |
| 2003-2019 |  | Prior to 2003 |
| Rural/Urban |  |  |
| Food insecurity |  | Non-environmentally affected livelihoods |
| Drought |  | Broad climate change focus and Future predictions |
| Africa |  | Beyond Africa |
| Environmental migration |  |  |
| ART |  | Children |
| Adherence |  | Thesis |
| Viral Load |  | Non-English publication |
| Human impacts (socio-ecological) |  | Non-socio-ecological impacts |

We imported all articles into Endnote reference management software (version X9, Thomson Reuters, Philadelphia, Penn.), and excluded duplicates using the ‘find duplicate’ function in Endnote. KO, SAK, DK, CI independently screened the titles and abstracts of all records to identify studies possibly related to our areas of interest. We obtained full texts articles from the three distinct searches (Table 1A) which examined the (i) general impact of drought on livelihoods (ii) drought on human health and (iii) those that examined HIV treatment adherence-related factors. KO and CI screened the full text articles based on the inclusion and exclusion criteria (Table 1B) and CI made final decisions on which article to be included in the review when there is a discrepancy. We included both Quantitative and Qualitative studies to allow us to describe both the proximal and distal factors that connect drought with HIV treatment adherence.

For the quality assessment of included articles, we applied the Critical Appraisal Skills Programme (CASP) quality assessment tool^16^ to specifically assess only studies linked directly to adherence as the outcome variable of interest. For Quantitative and Mixed-methods studies, the CASP^16^ criteria we addressed the following questions: (i) Is the question clear or are there clear aims and objectives? (ii) Is the sample appropriate, and does the size allow generalisation? (iii) Is the research design clearly stated? (iv) Is the data collection process clear including recruitment and consent? (v) Did the researcher follow the steps of data analysis and was the data management clear? (vi) Are the results accurate and presented in the correct format? (vii) Does the discussion and conclusion support the results?

To assess the quality of Qualitative studies, a previously described adaptation of questions representing the three key conceptual domains described in the CASP quality assessment tool was used.^16,17^ The criteria addressed the following questions: (i) Was the relationship between researcher and participant adequately considered; (ii) Was the sampling method clearly described; (iii) Was the data collected in a way that addressed the research issue; and(iv) was the analysis method clearly described?

We organised the studies by year of publication, study design, country of origin, and key findings. For each search, we grouped findings into key thematic areas using NVivo 12 Pro (QSR International) and Microsoft Excel to tabulate them. Subsequently, we linked common themes across the searches to establish the relationships between drought, health, HIV and adherence to HIV care and treatment using the same approach for both Quantitative and Qualitative studies. We did not undertake a meta-analysis in this review.

## 3. Results: A systems understanding of the sensitivity of ART adherence to factors linked to drought

The number of articles derived from the search of the databases are summarised in Figure 1. Our search identified 3217 articles, and we excluded 503 duplicates. A further 2482 were excluded after screening abstracts for titles and relevance, leaving 232 articles relevant for full text review, of which 121 articles were excluded as they did not meet inclusion criteria on adherence as the main outcome variable of the study, situated in Africa, focused on adults, and human impacts in the case of drought (Table 1B). Studies that focused broadly on climate change without direct emphasis on drought or those that mention adherence without specific focus on this outcome and related factors were adjudged insufficient and consequently excluded (Figure 1). Finally, after all exclusions 111 articles were synthesised in the systematic review, including 71 quantitative studies, 27 mixed methods and 13 qualitative studies (Tables 2A & 2B).

**Figure 1:**
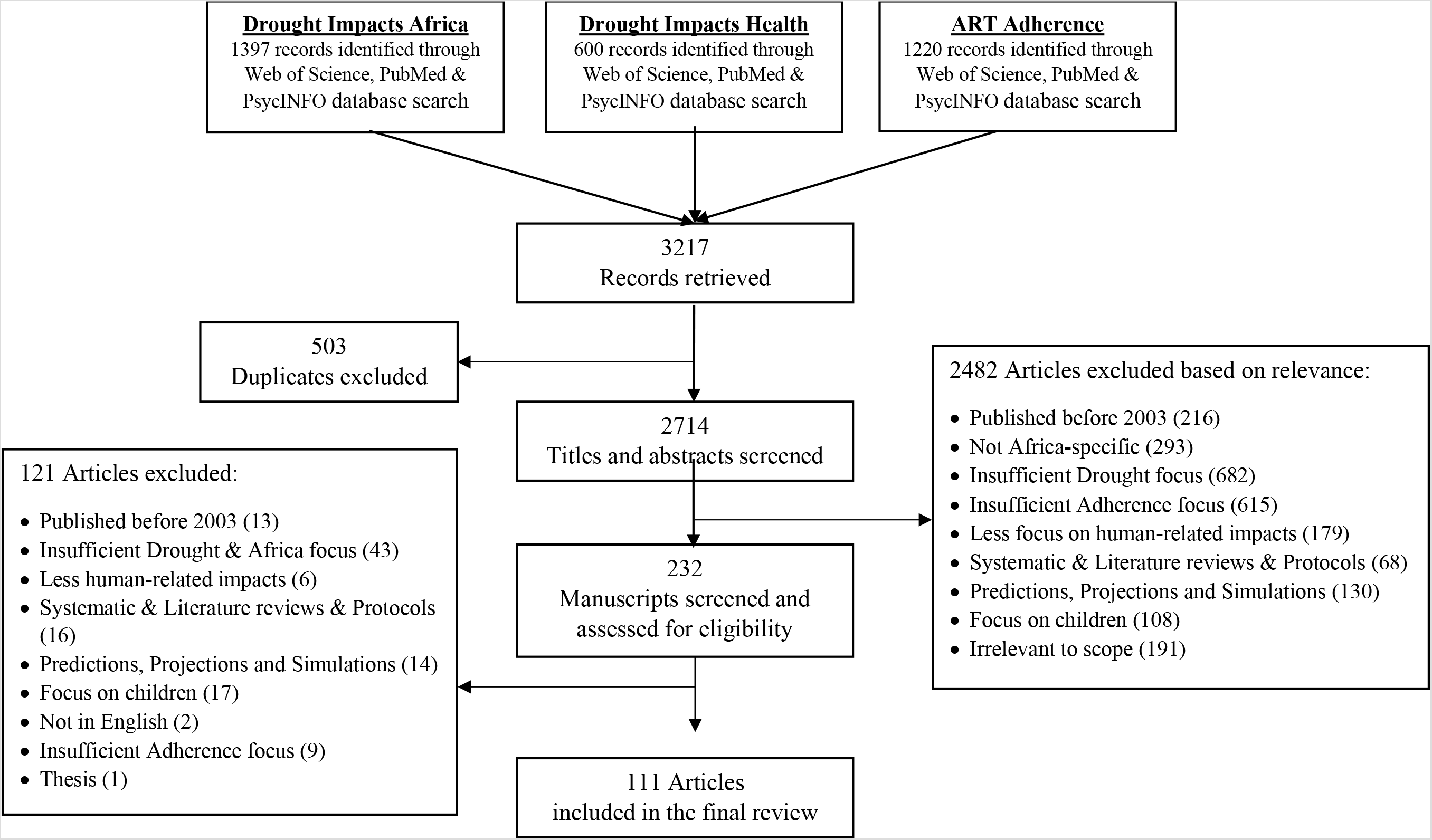
Flow chart of search results and included studies.

**Table 2.** Reviewed studies by methods and socio-ecological level.

| **A: All reviewed Studies (n=111)** |  |  |  |  |
| --- | --- | --- | --- | --- |
|  | **Drought impact** | **Drought on Health** | **ART Adherence** |  |
| **Quantitative studies** | 33 | 9 | 29 | 71 |
| **Mixed methods** | 22 | 3 | 2 | 27 |
| **Qualitative studies** | 5 | 1 | 7 | 13 |
|  | 60 | 13 | 38 | 111 |
| **B: Reviewed Adherence-related Studies (n=38)** |  |  |  |  |
|  | **Quantitative** | **Mixed methods** | **Qualitative** |  |
| **Individual level** | 23 | 2 | 6 | 31 |
| **Community/contextual level** | 13 | 2 | 5 | 20 |
| **Health systems/policy level** | 11 | 1 | 2 | 14 |

Many studies exploring factors associated with ART adherence have relied on a socioecological framework. This framework recognises that societal norms and structures influence individual attitudes and behaviours, and identifies key levels impacting adherence to ART: individual (knowledge, attitudes, beliefs, perceptions), community (cultural values and norms), interpersonal (family, friends, social networks), Institutional (health system, social institutions, work place) and public policy (local, state and national laws and policies).^18–20^ The most popular of these levels that we used to delineate adherence-related factors include individual, community or contextual, and health or policy systems levels (Table 2B). We note that of 38 adherence-related studies, 31 highlighted individual level factors (23 quantitative, three mixed-methods, five qualitative studies). 20 studies (13 quantitative, two mixed-methods and five qualitative studies) addressed community and contextual level factors. Health systems factors were addressed in 11 quantitative studies, one mixed-methods and two qualitative studies. The quality assessment result showed that only five of the 31 included quantitative and mixed-methods studies were deemed to be at low risk of bias (Supp. Table 2) while only one of the seven included qualitative studies was found to be at low risk of bias (Supp. Table 3). All 111 studies that contributed to the synthesis are summarised in Table 3.

**Table 3.** Thematic Areas showing Interlinked factors between Drought and ART (non)Adherence*.

| **Livelihoods and Economic conditions (total studies=76)** | **N (%) studies** | **Social Support and Psycho-Behavioural disposition (37)** | **N (%)** | **Comorbidities and ART Regimens (27)** | **N (%)** | **Human Mobility (17)** | **N (%)** |
| --- | --- | --- | --- | --- | --- | --- | --- |
| Catastrophic treatment costs | 2 (3%) | Age | 4 (10%) | Medication side effects | 11 (40%) | Migration and Displacement | 14 (82%) |
| Drought Impact Mitigation | 23 (30%) | Alcohol and Substance abuse | 6 (16%) | Nurses and officials' behaviour | 4 (14%) | Seeking off-farm employment | 2 (12%) |
| Food insecurity | 22 (29%) | Fertility choice | 6 (16%) | Drought-related diseases | 2 (7%) | Travel away from but not migration | 3 (18%) |
| Poor ART knowledge | 4 (5%) | Inadequate counselling or support groups | 6 (16%) | Comorbid conditions | 16 (59%) |  |  |
| Intervention programs | 2 (3%) | Depression and mental ill-health | 5 (14%) | Stock-out | 2 (7%) |  |  |
| Loss of production | 20 (26%) | Forgetfulness | 3 (8%) | Time on ART/Fatigue | 4 (14%) |  |  |
| Low water quality and quantity | 5 (7%) | Gender | 4 (10%) |  |  |  |  |
| Missing Education | 4 (5%) | Lack of CHW and LHW | 1 (3%) |  |  |  |  |
| Poor Individual and National Economic conditions | 5 (7%) | Less equitable gender norms | 2 (5%) |  |  |  |  |
| Reduced livelihood diversification | 2 (3%) | Marital status | 2 (5%) |  |  |  |  |
| Selling off assets and borrowing | 6 (8%) | Nondisclosure | 7 (19%) |  |  |  |  |
| Technology | 1 (1%) | Risky sexual behaviour | 2 (5%) |  |  |  |  |
| Unemployment | 15 (20%) | Stigma | 6 (16%) |  |  |  |  |
|  |  | Traditional belief and treatment | 2 (5%) |  |  |  |  |
|  |  | Violence | 8 (21%) |  |  |  |  |

*\*Note: The numbers shown in the column headings do not represent the cumulative numbers from individual factors within each theme, but the total number of unique studies in which the factors were mentioned; CHW = community health workers; LHW = Lay health workers.*

**Table 3.**
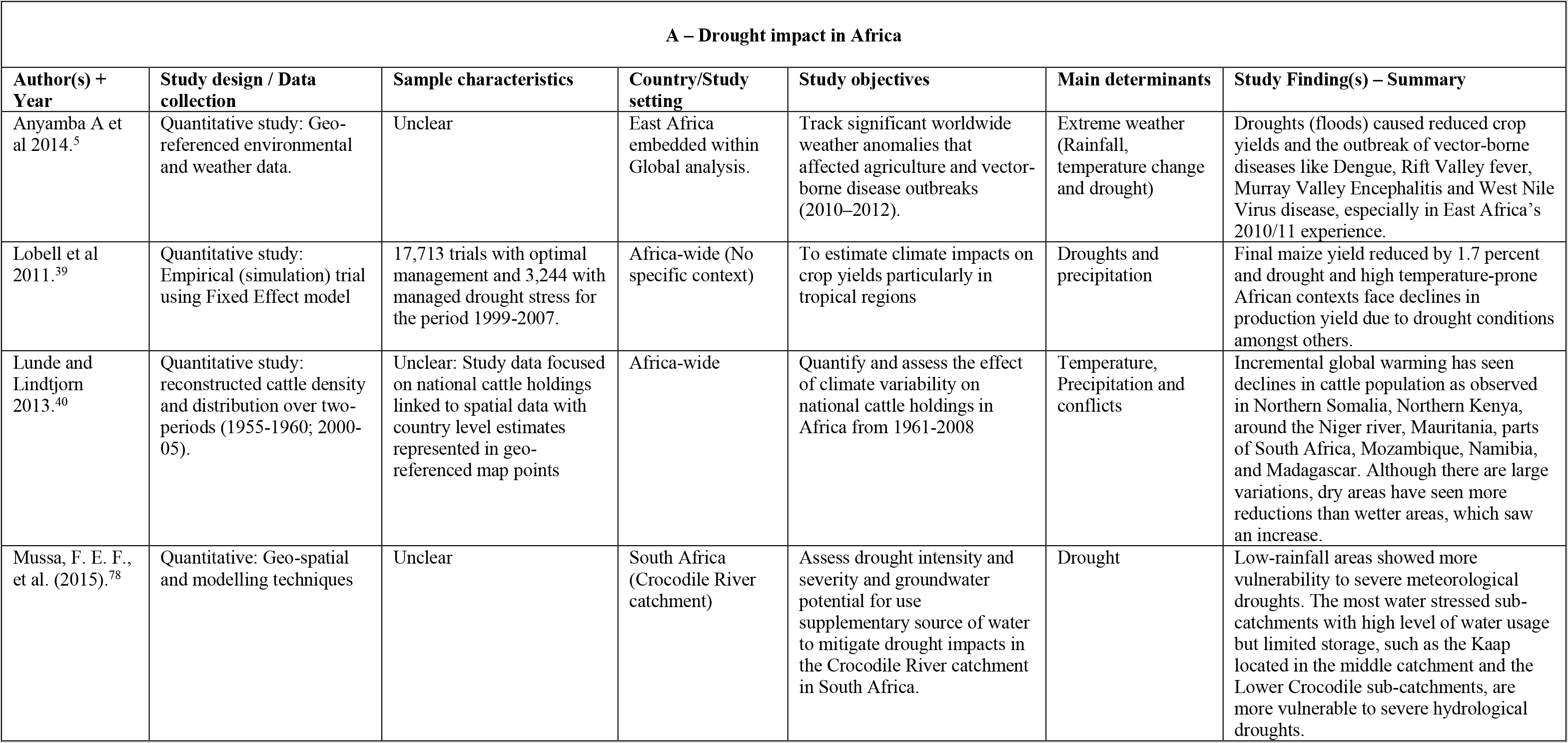
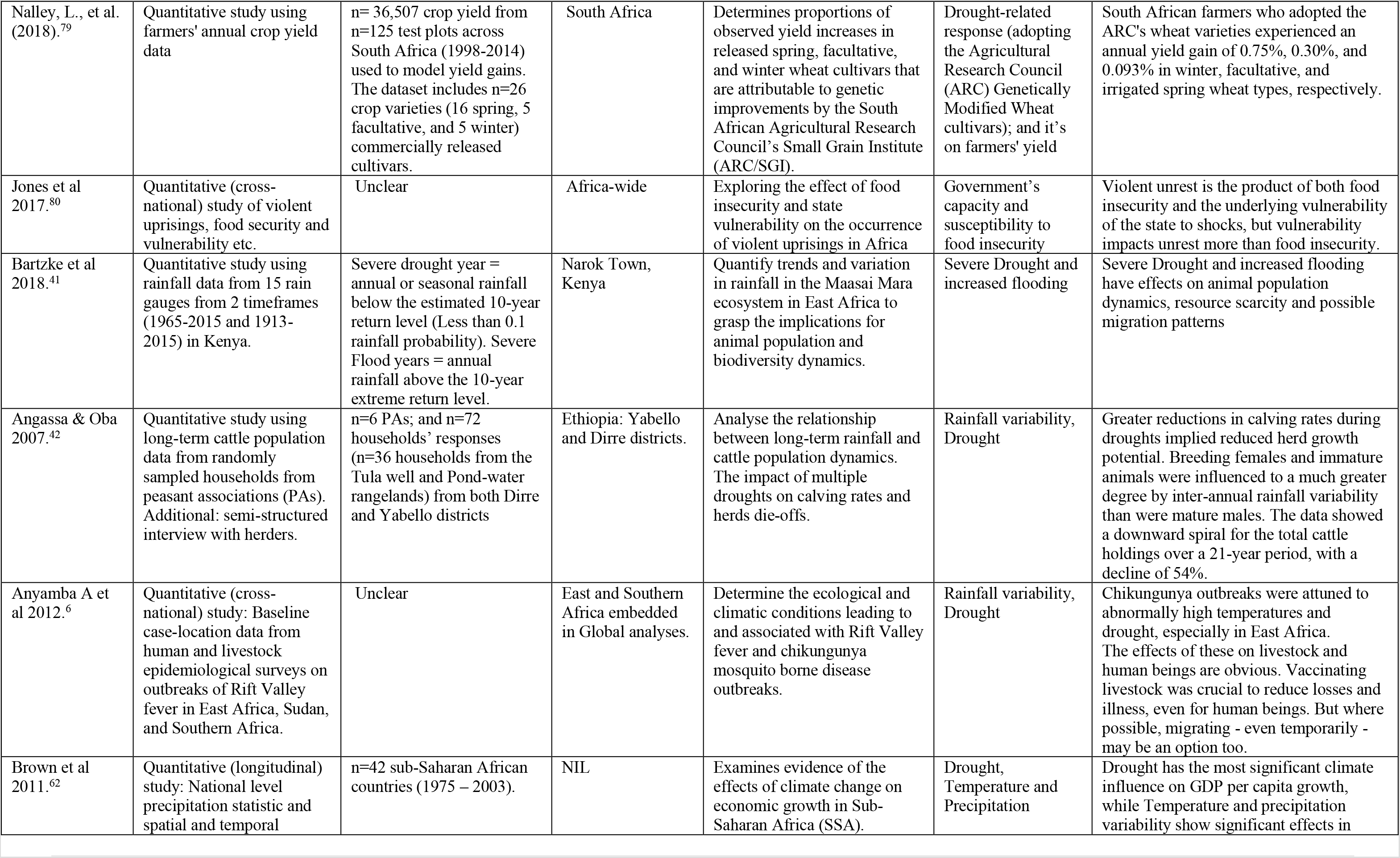
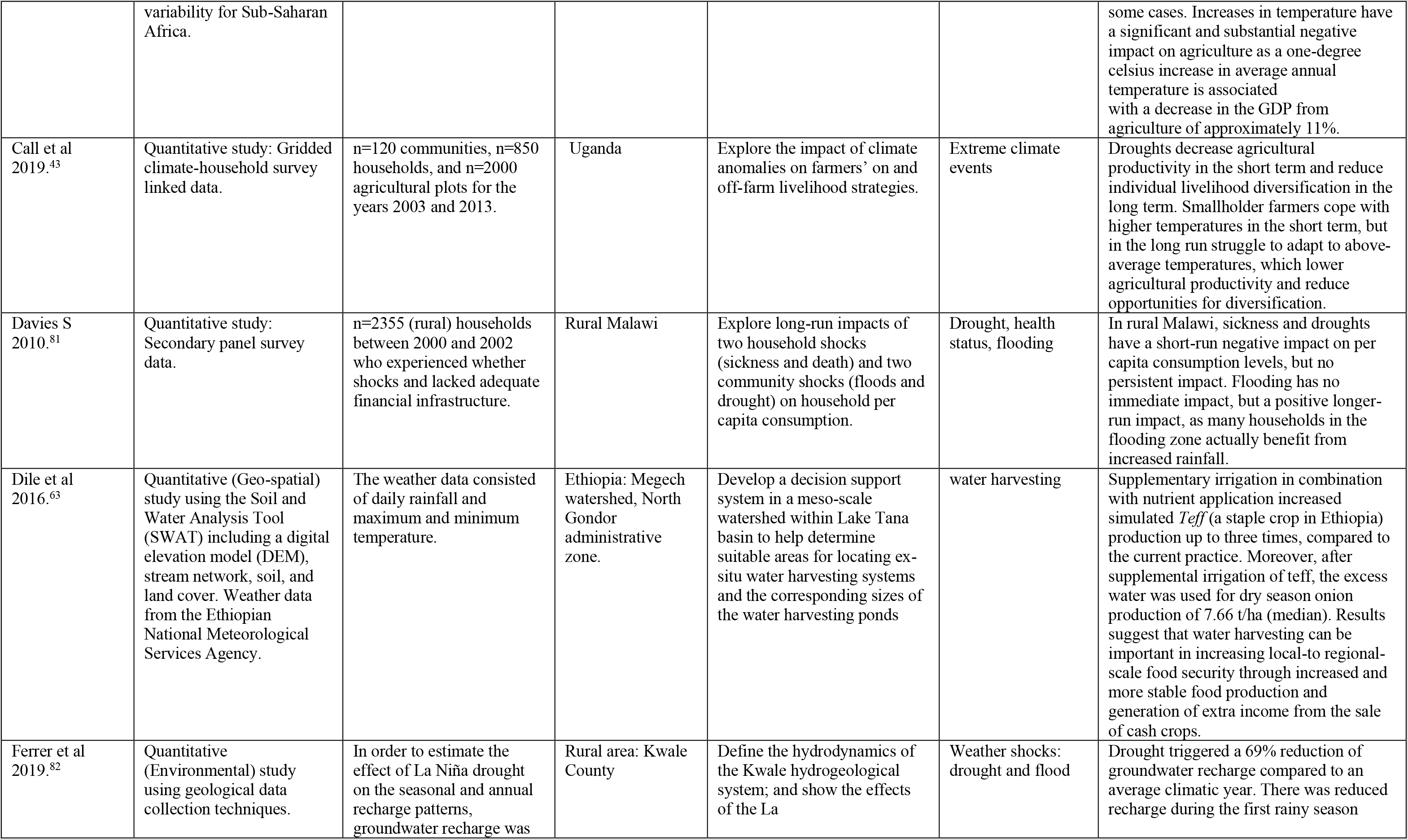
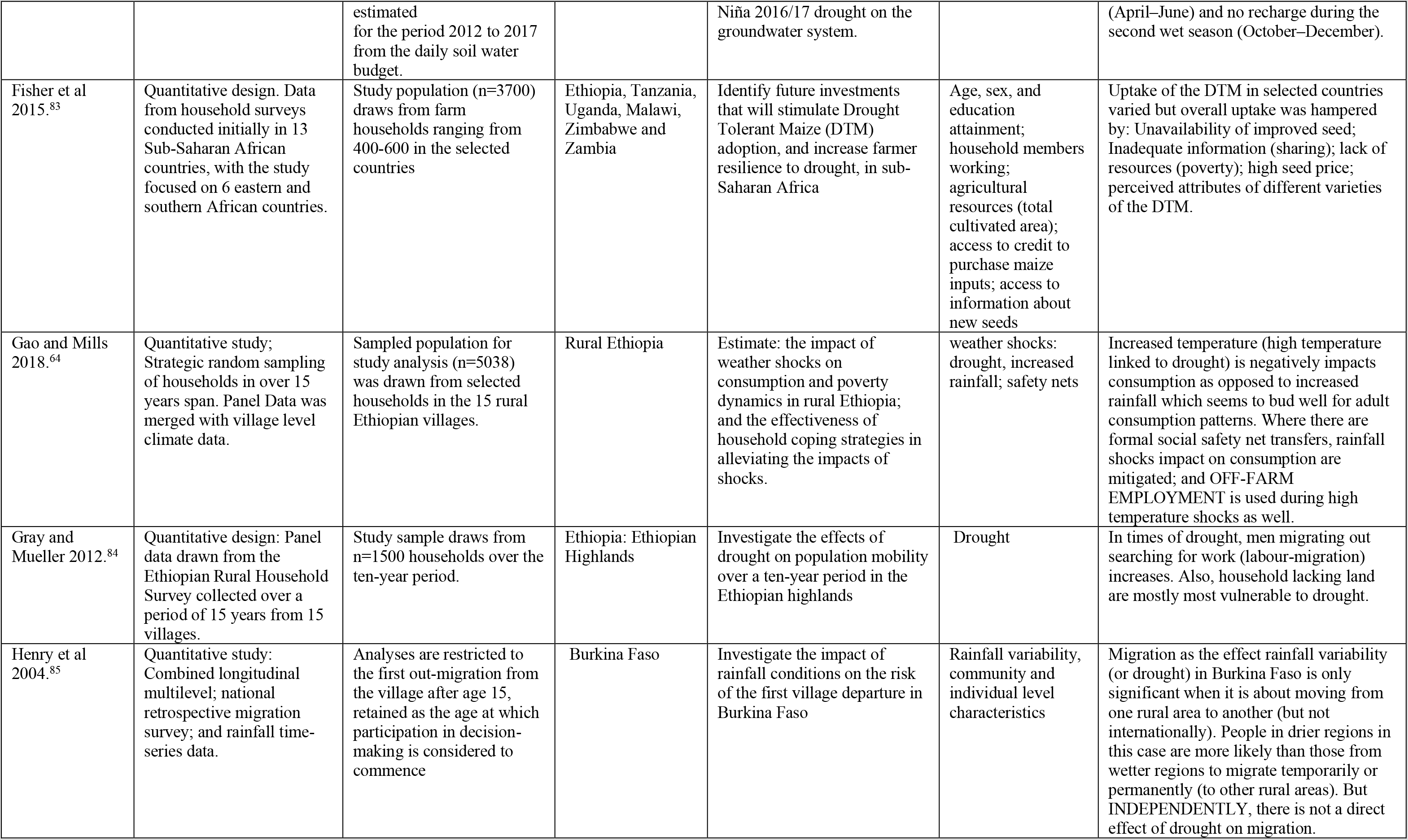
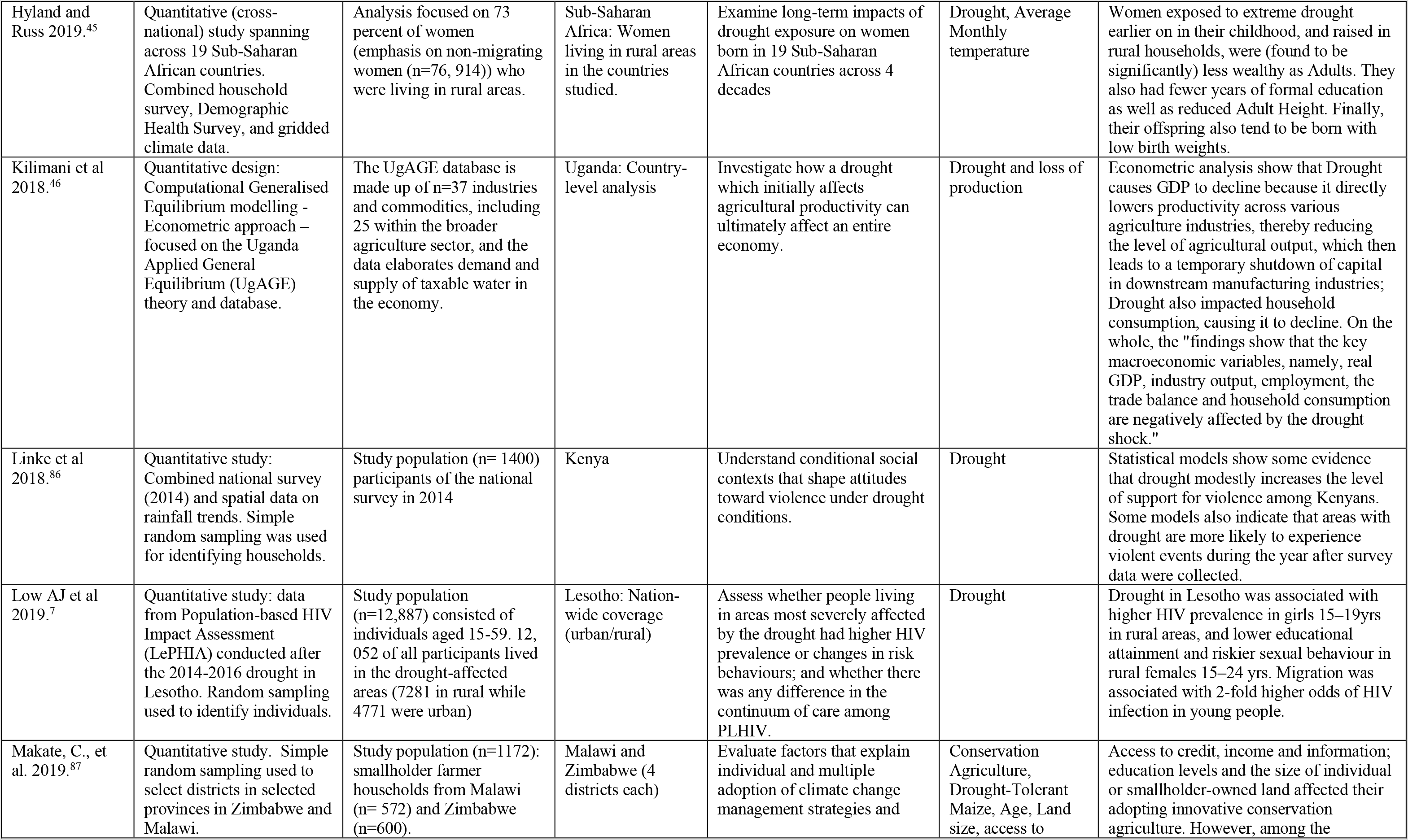
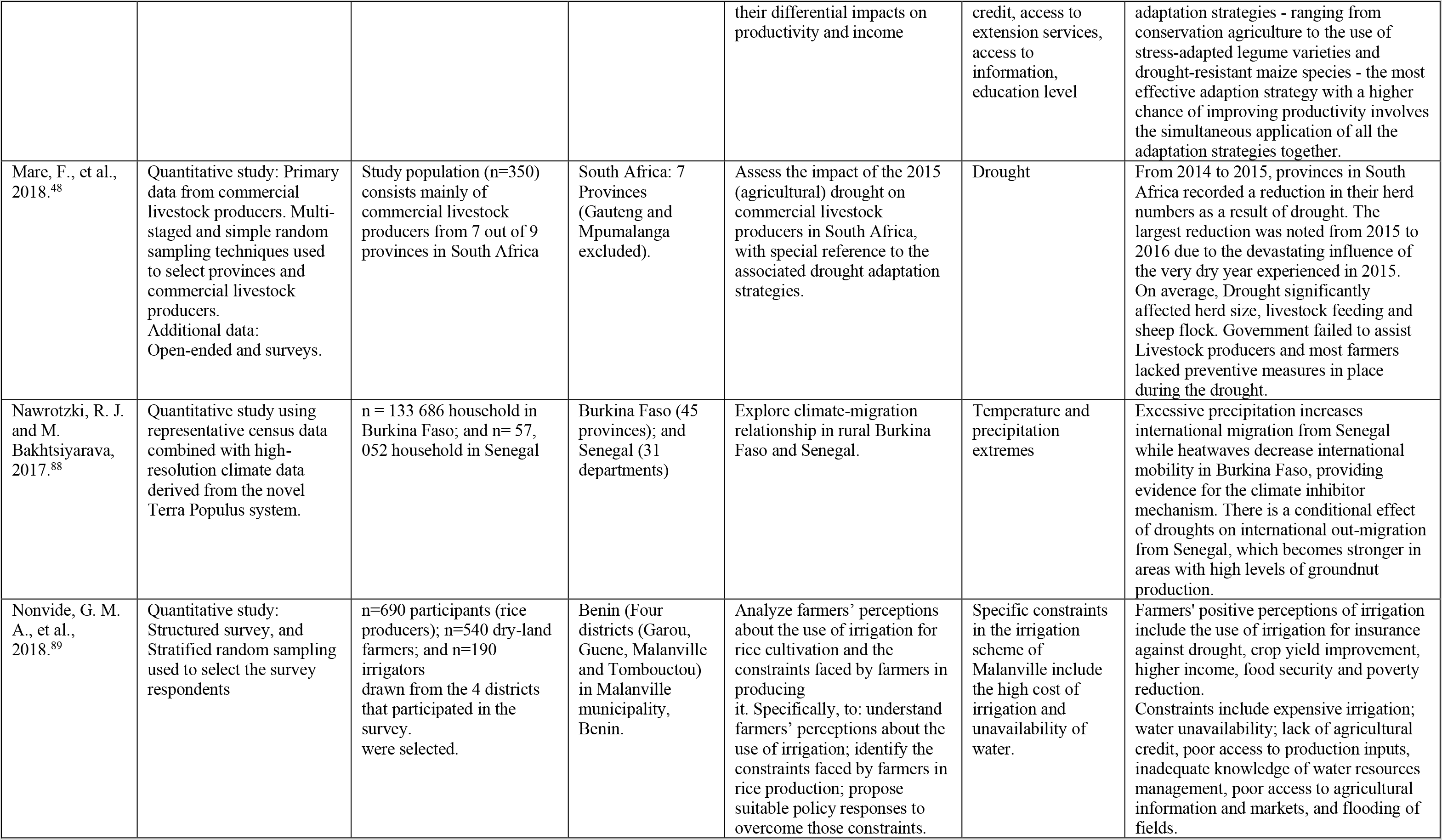
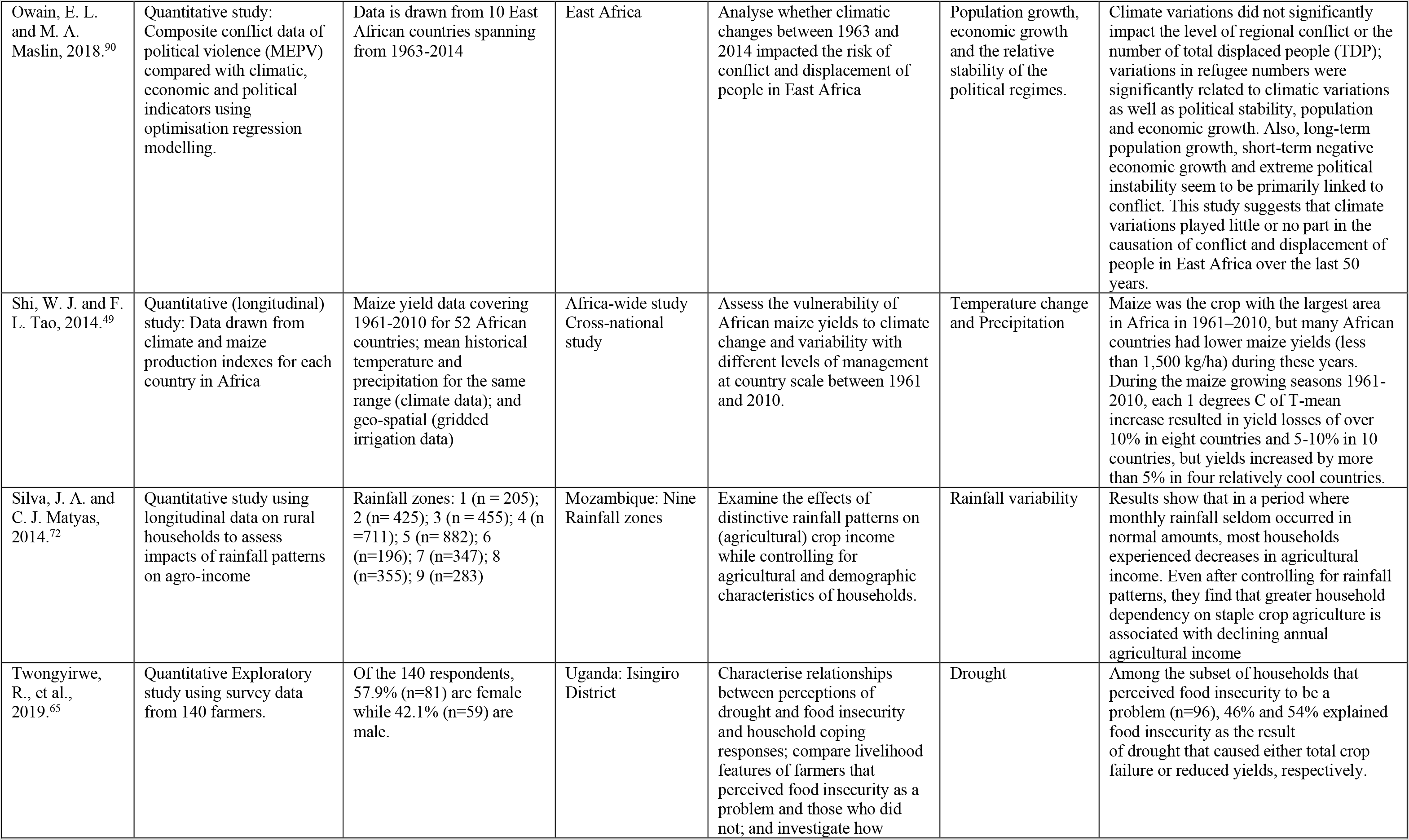
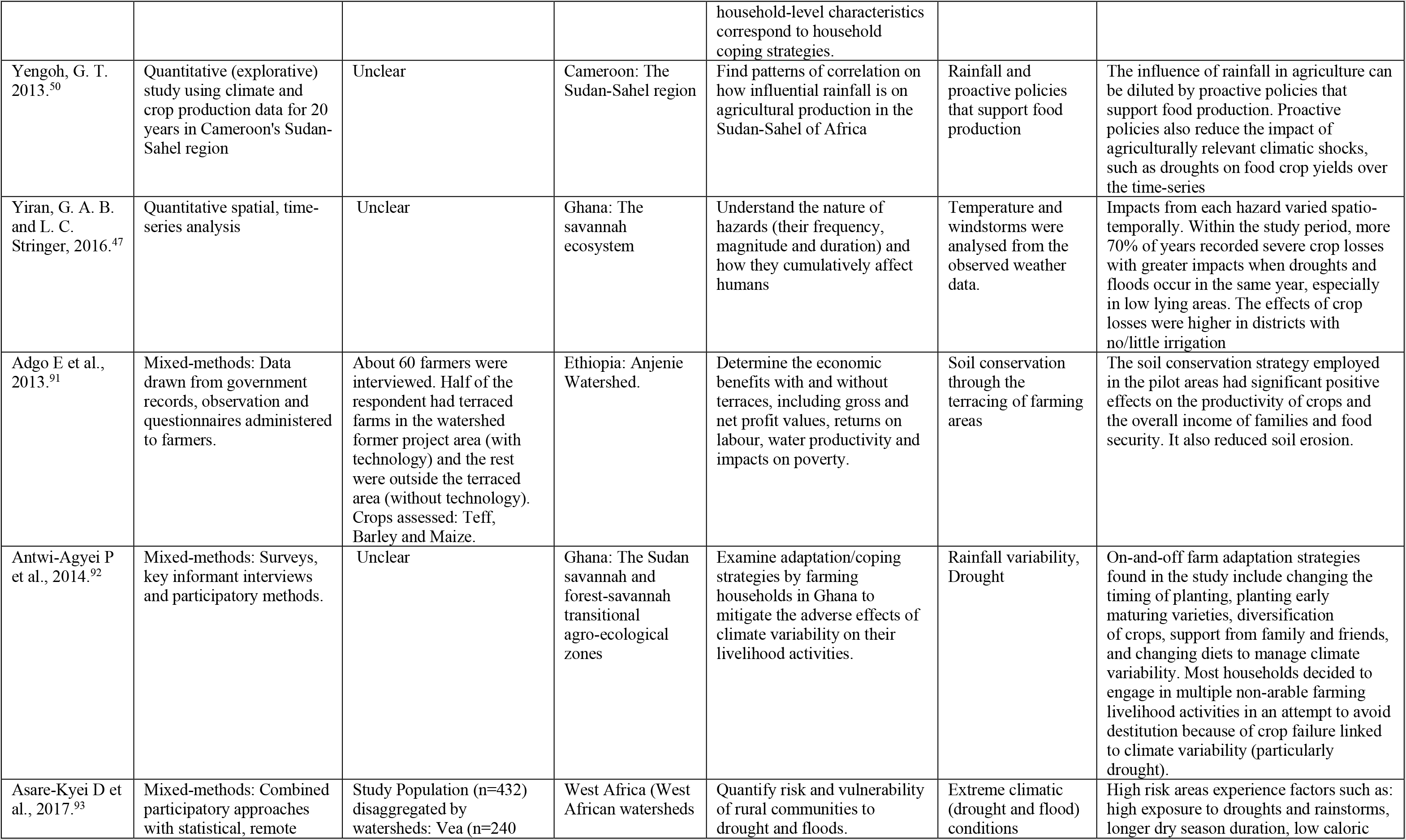
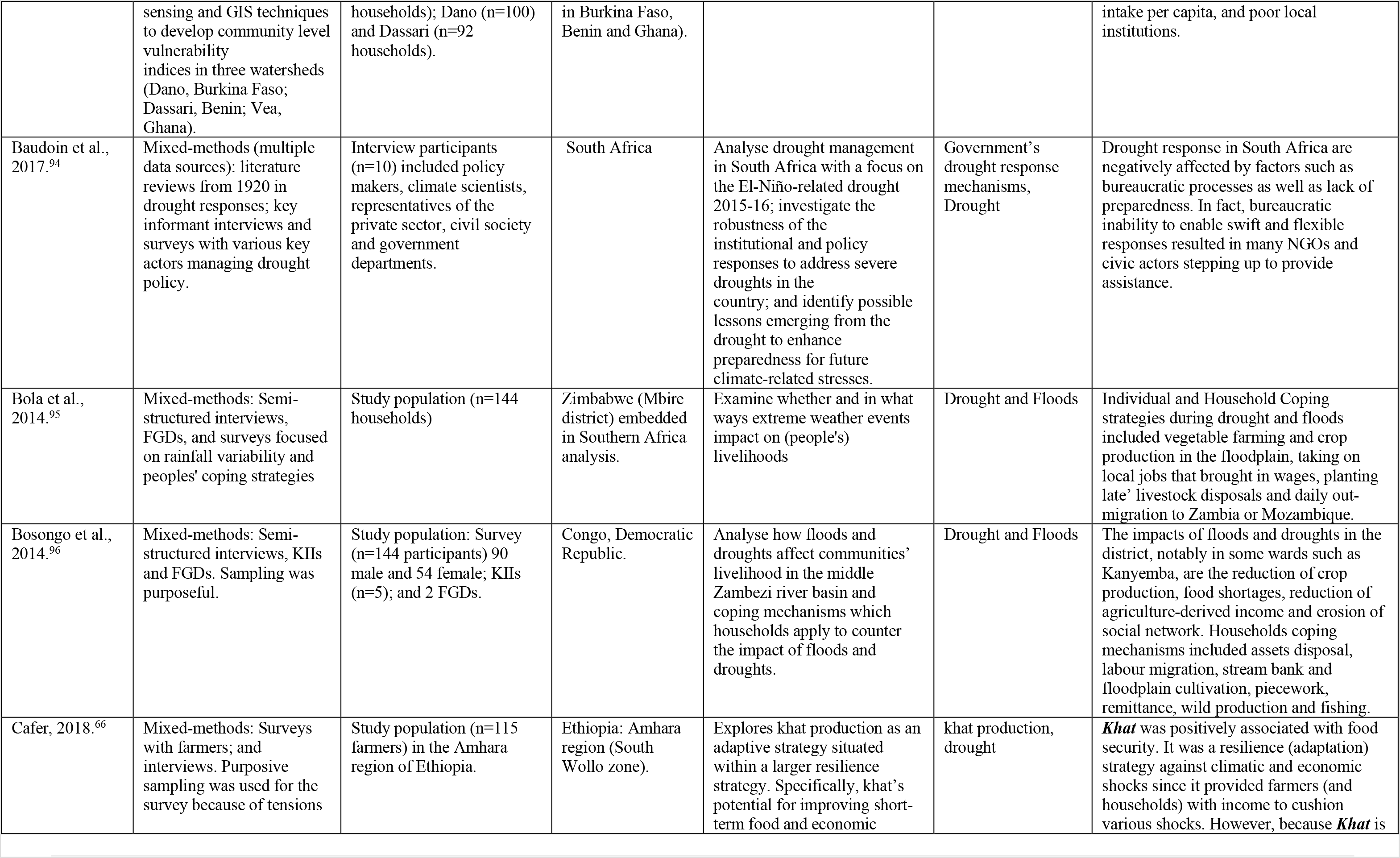
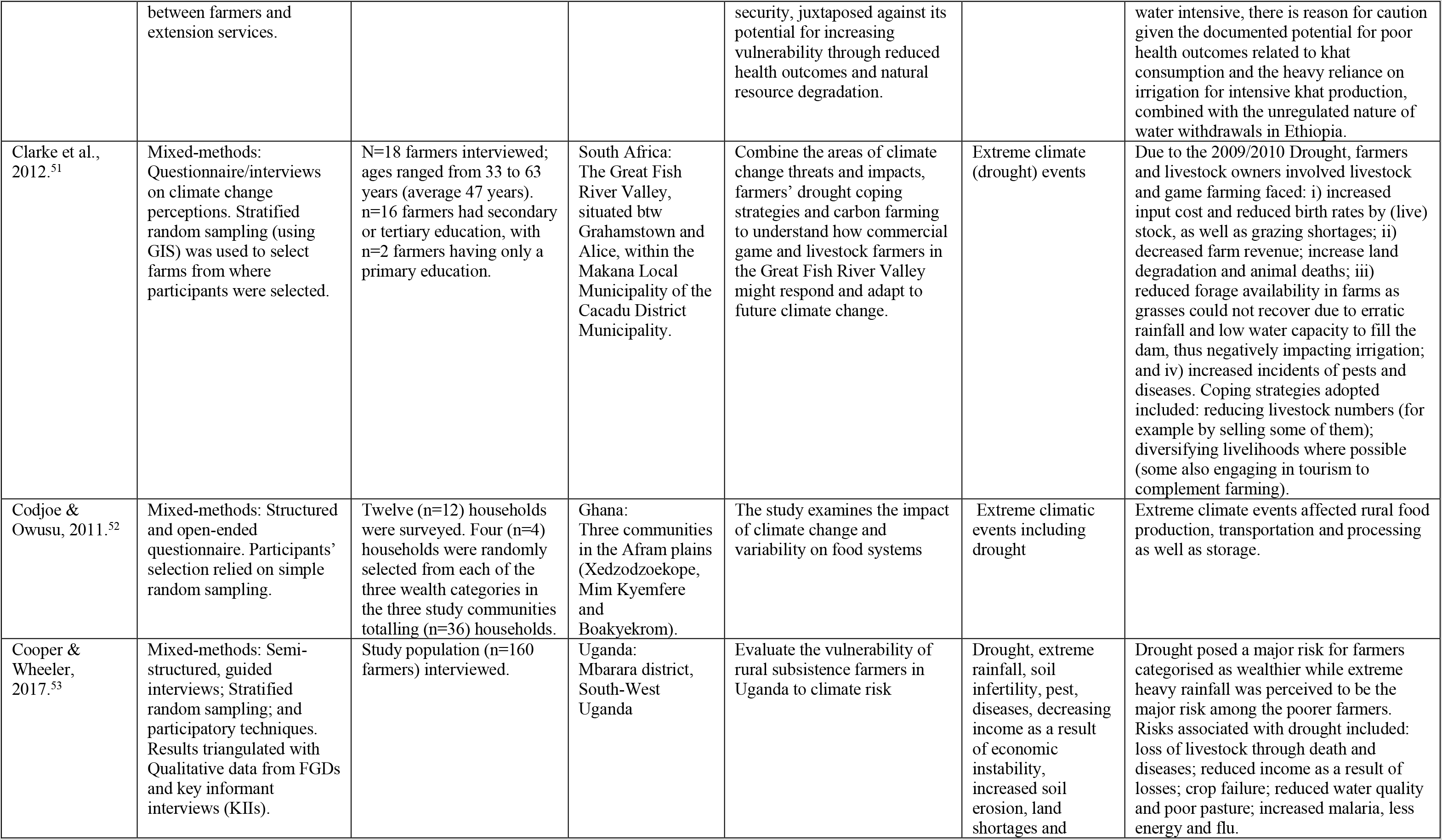
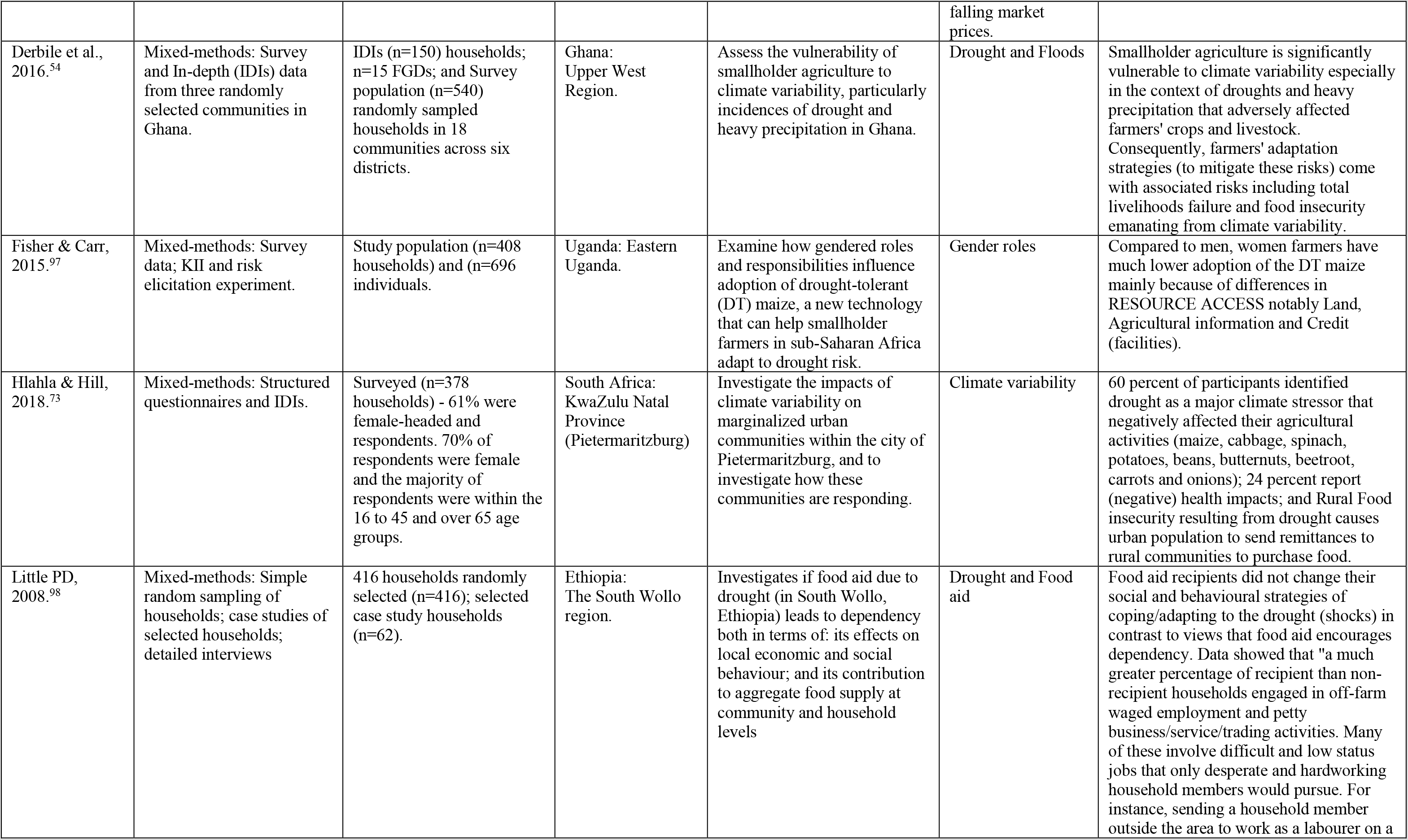
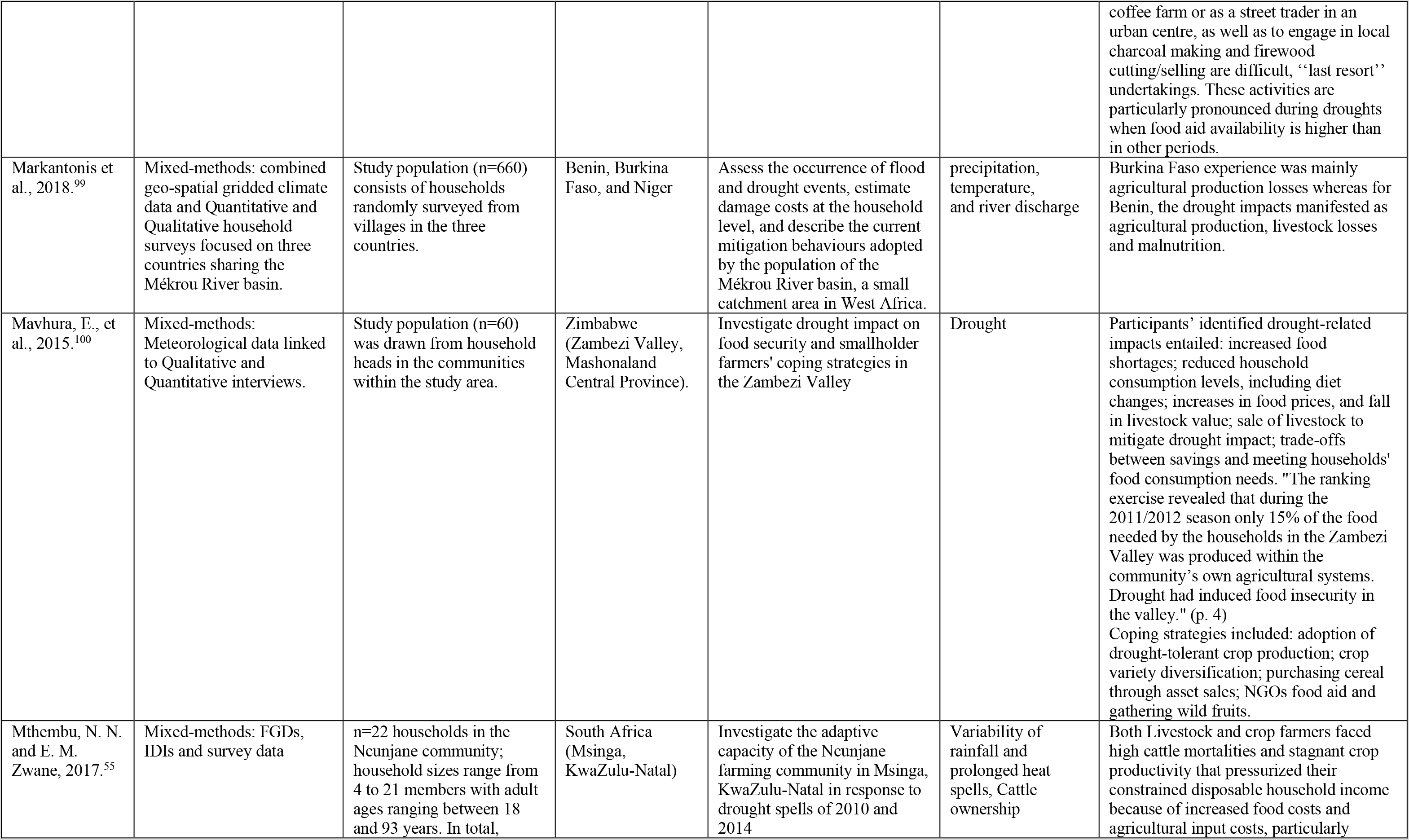
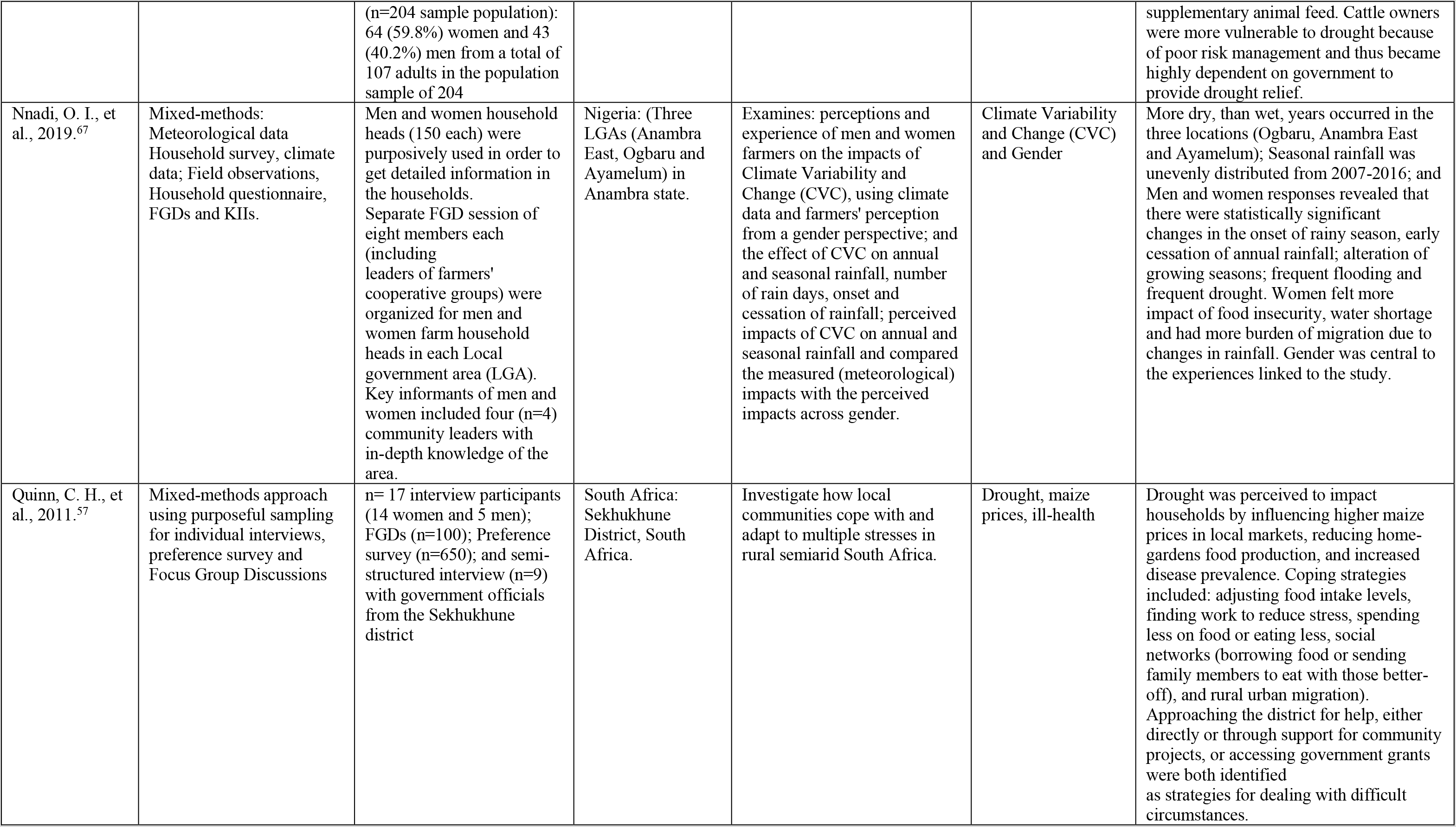
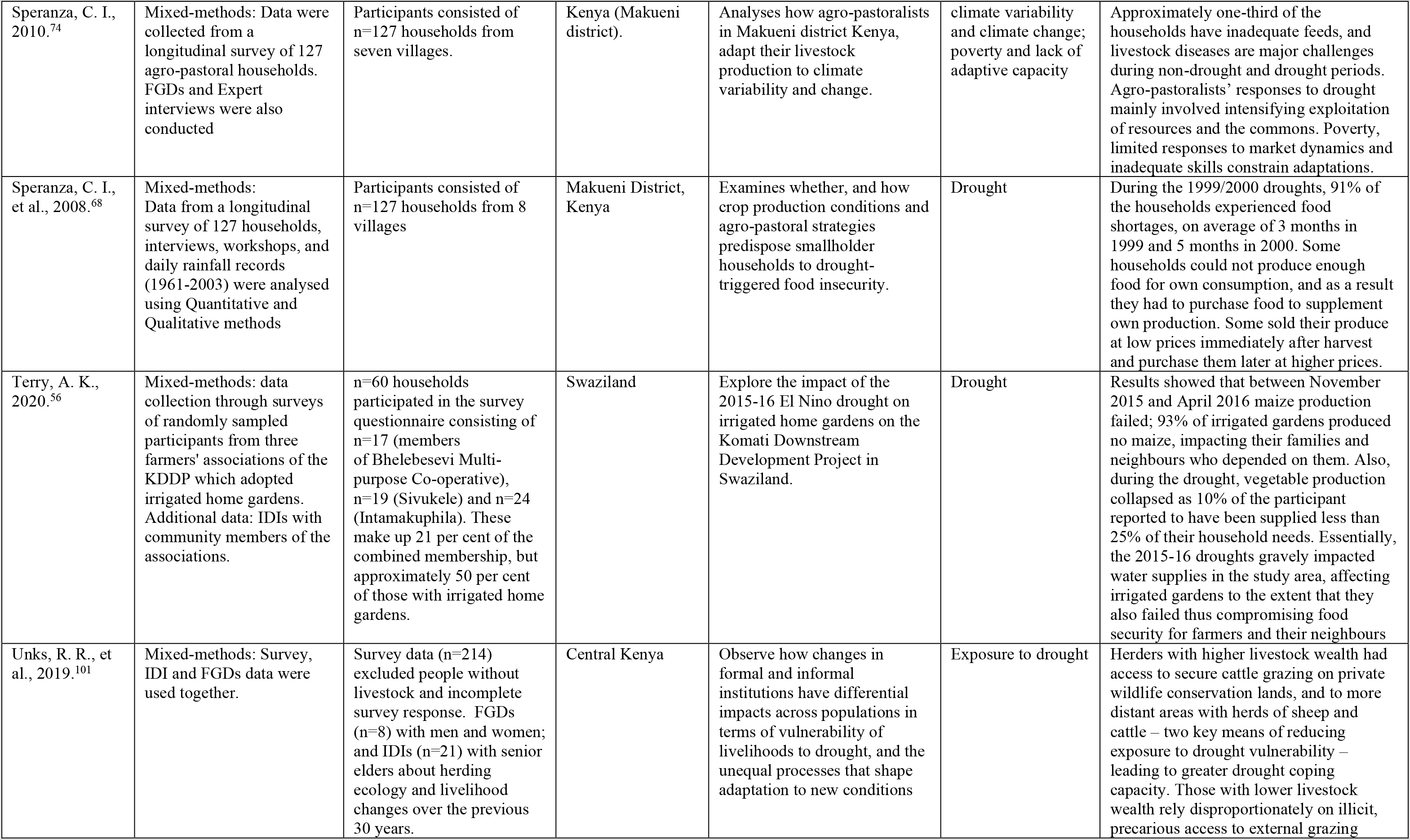
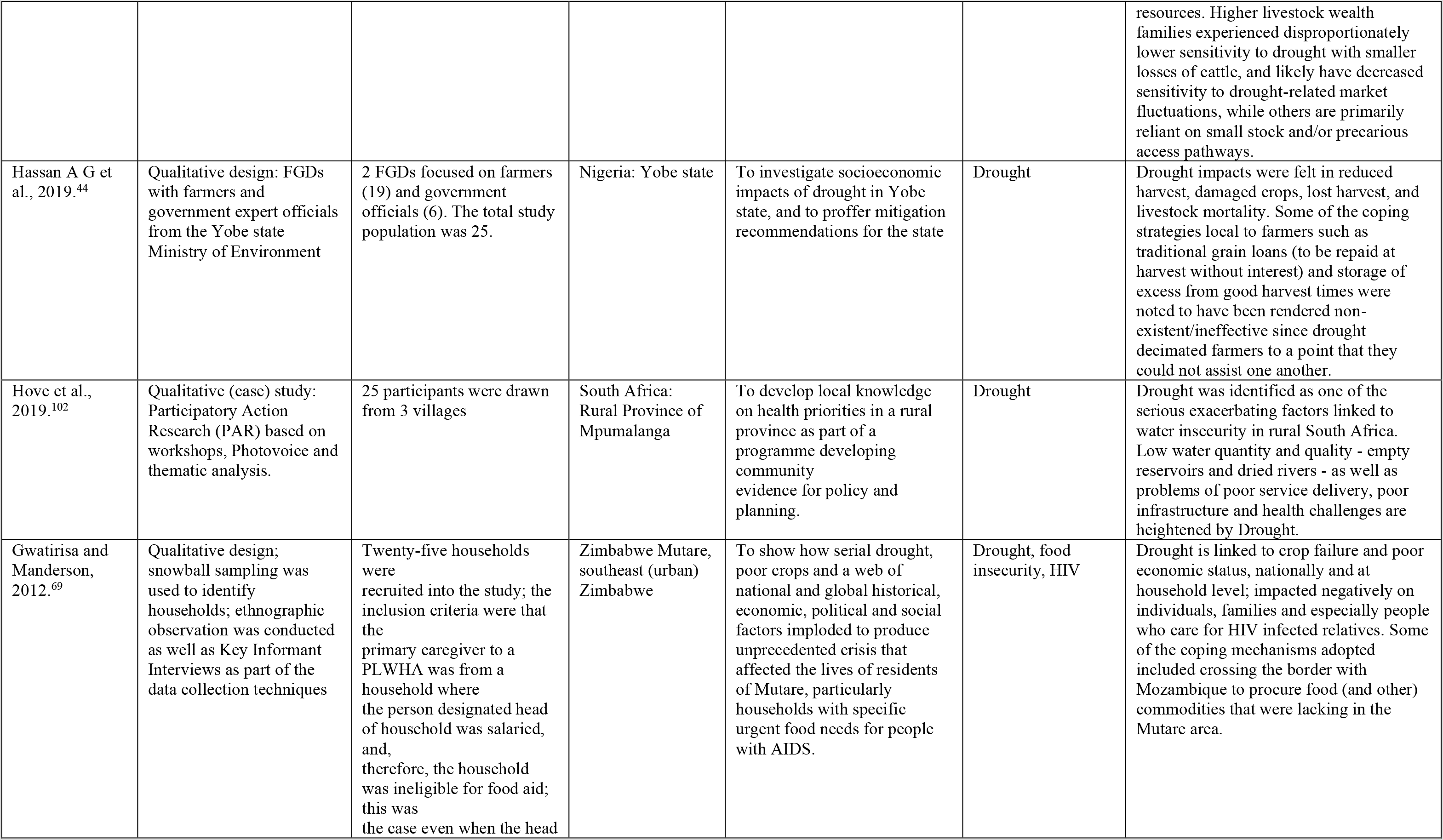
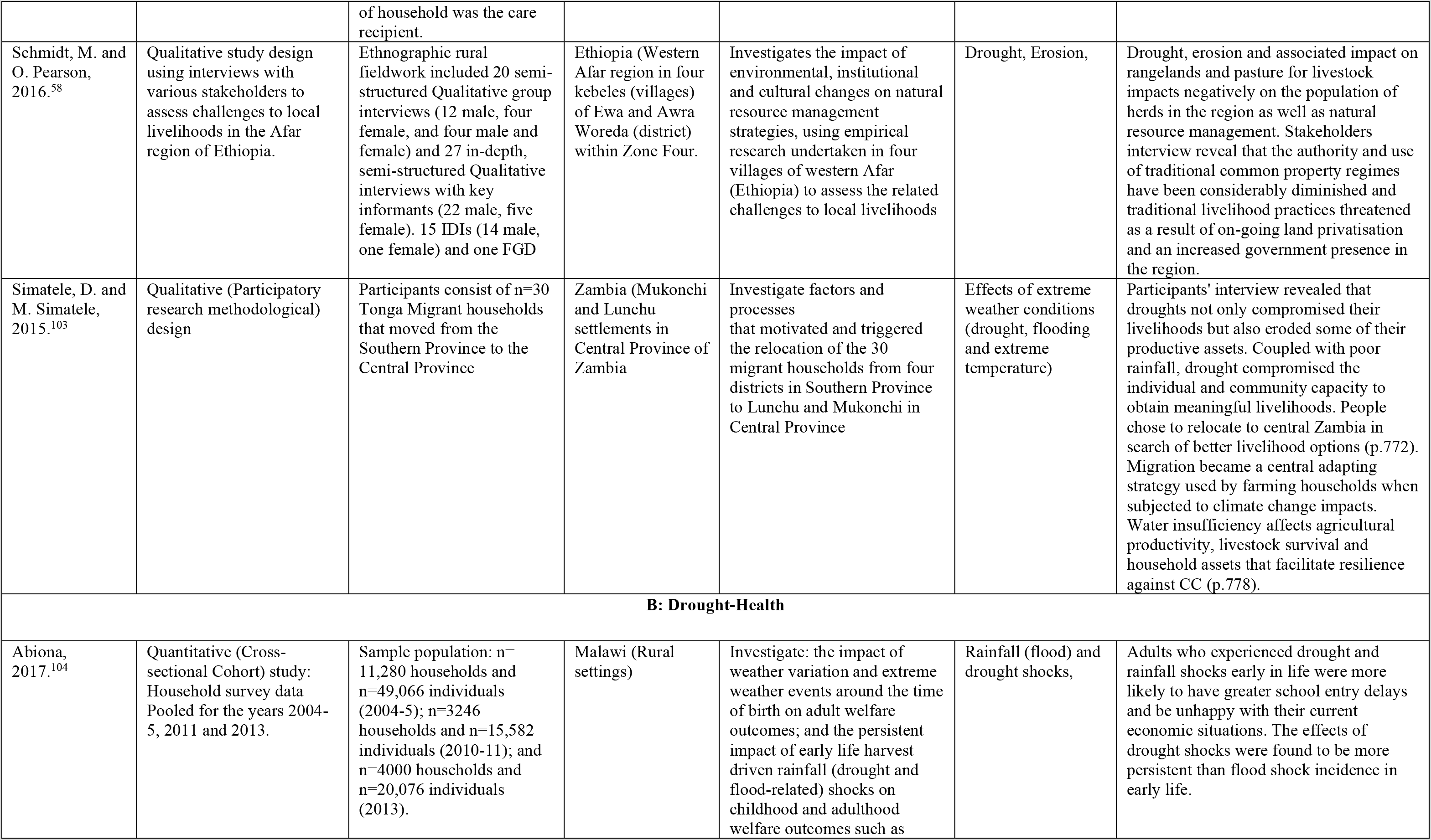
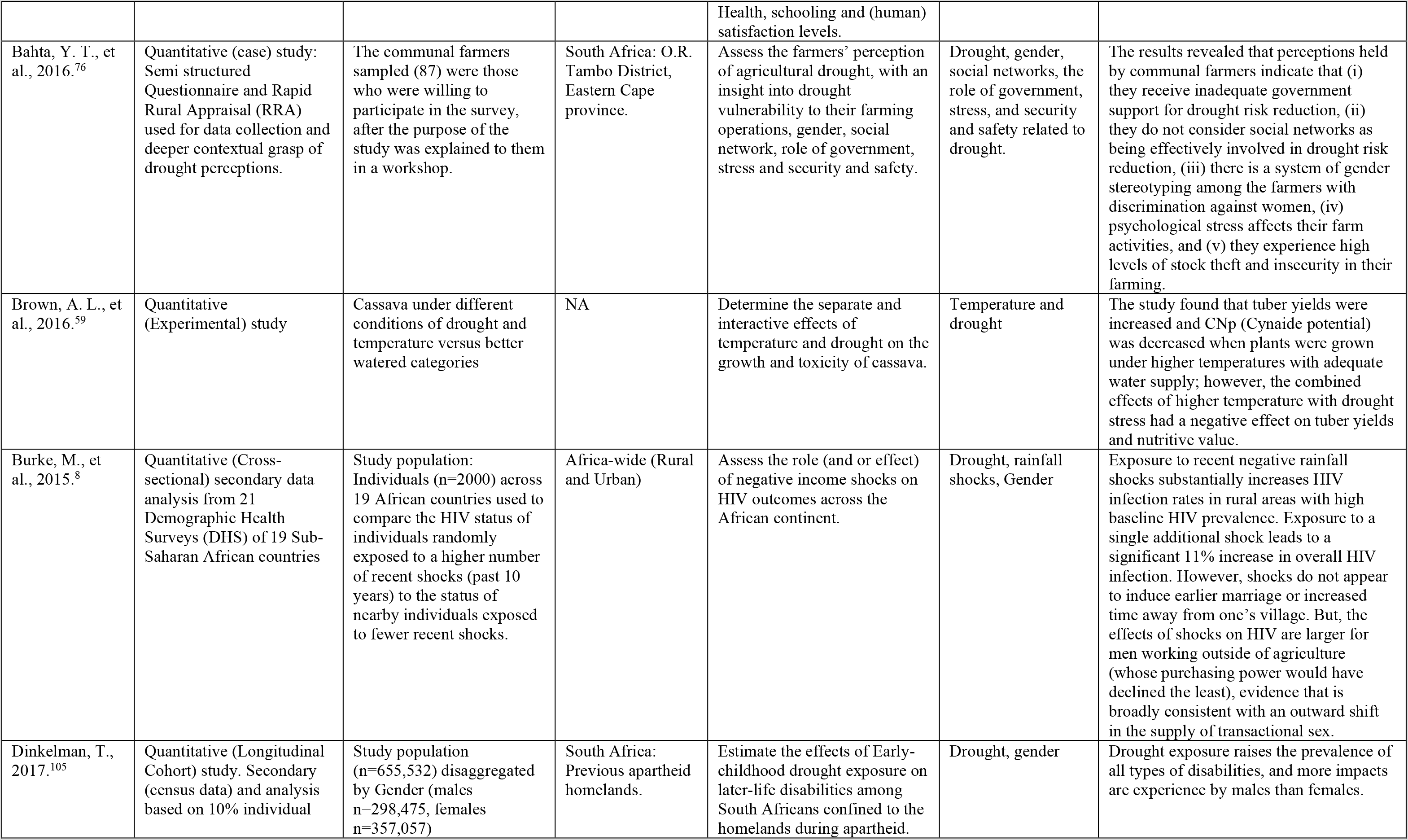
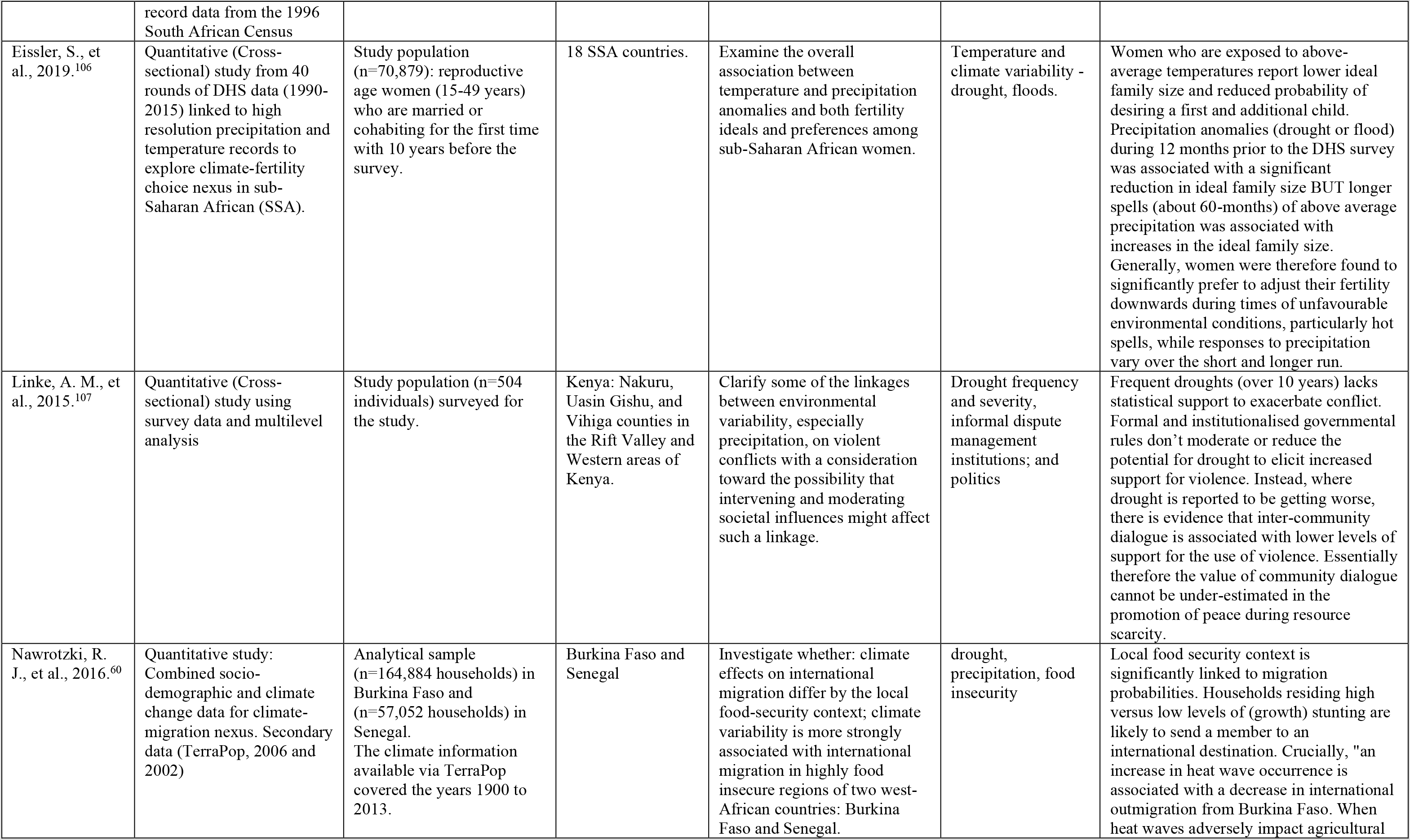
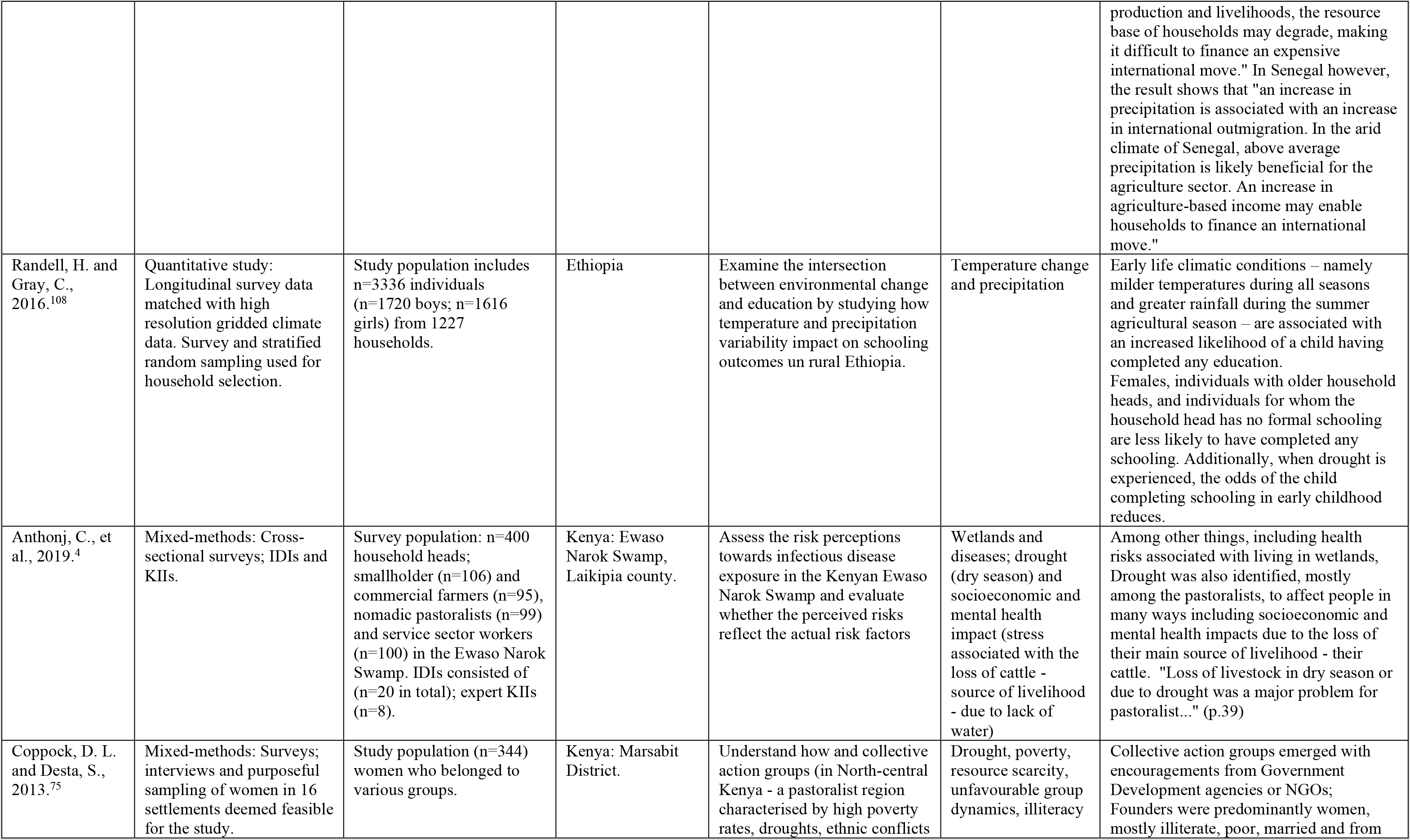
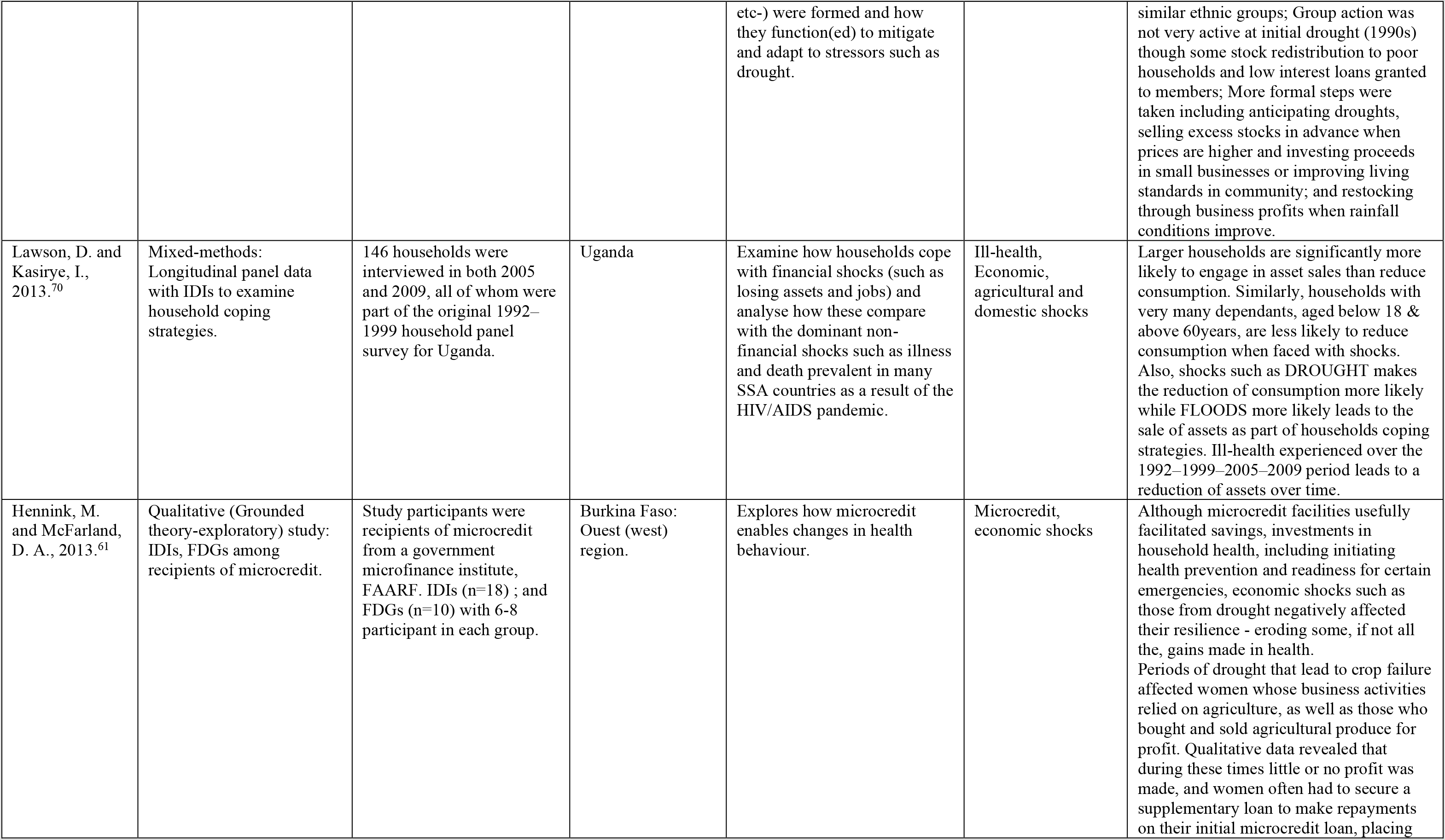
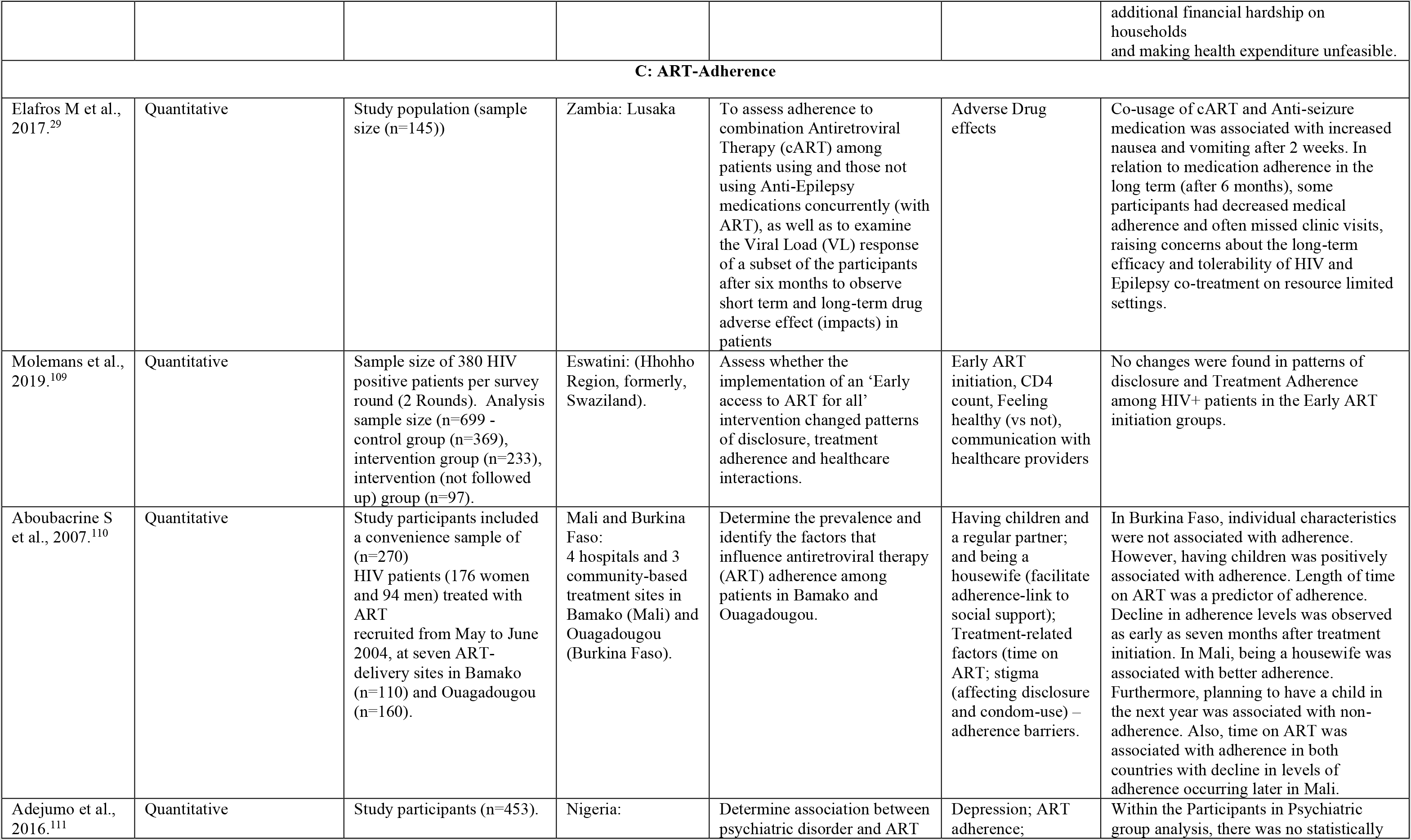
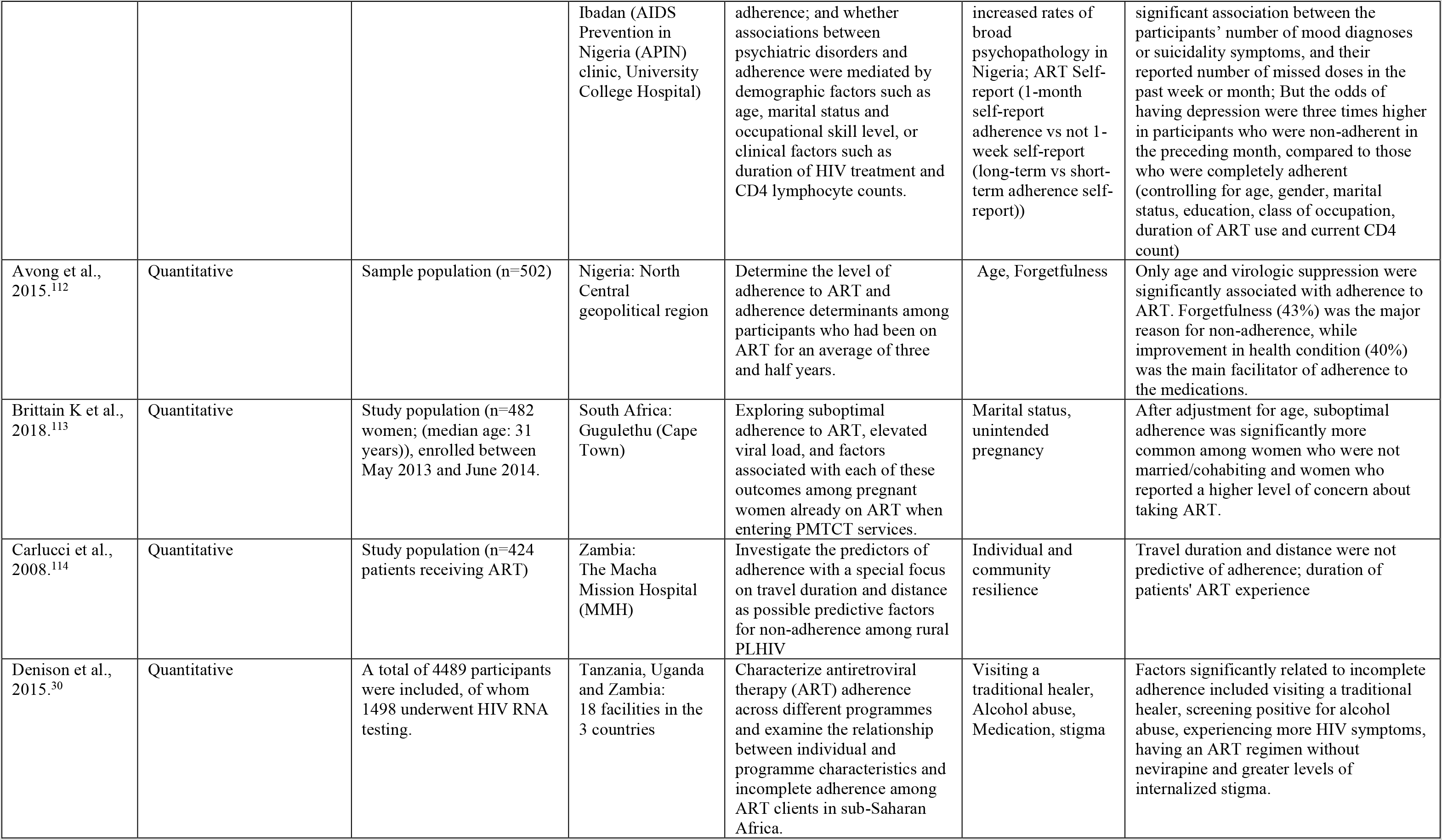
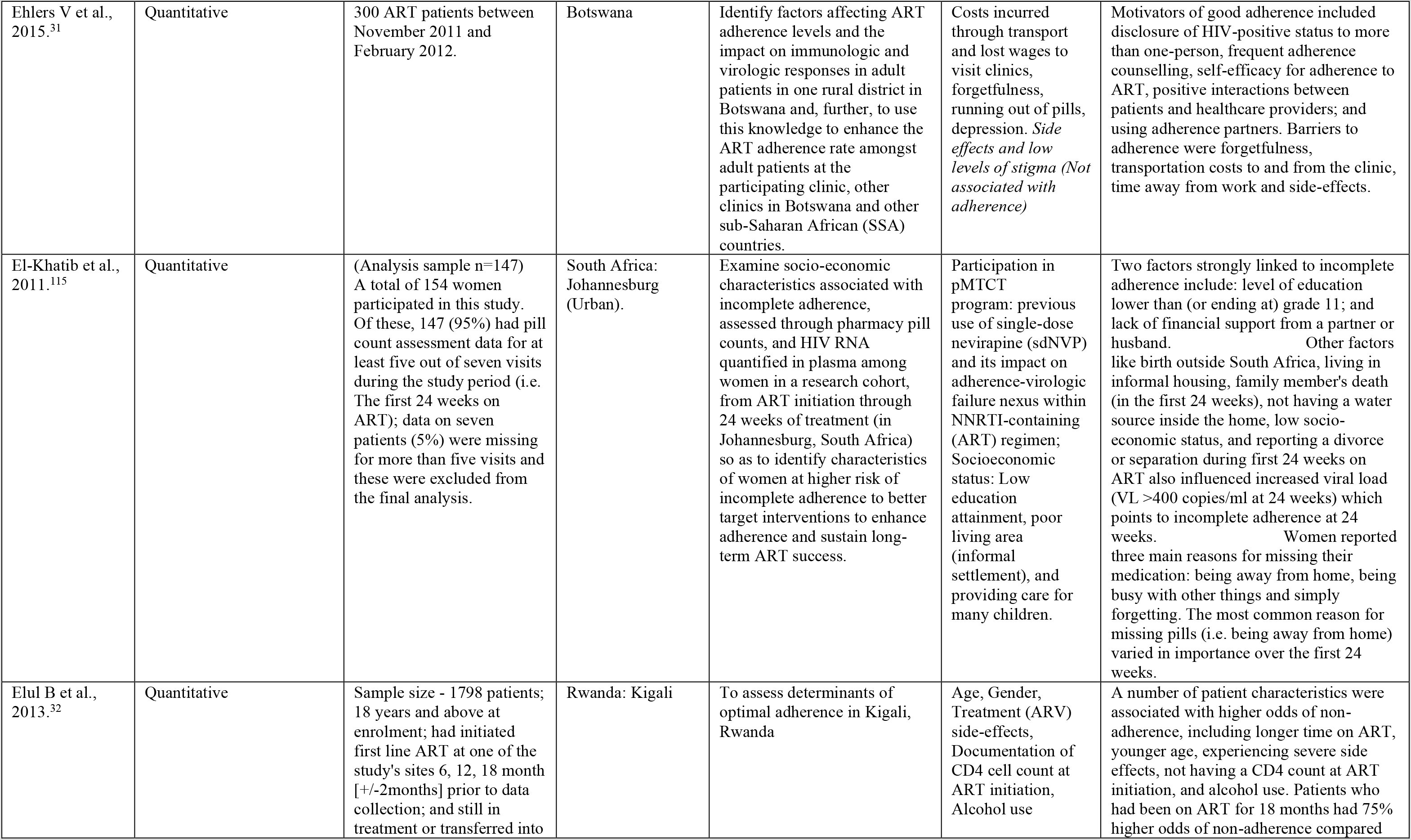
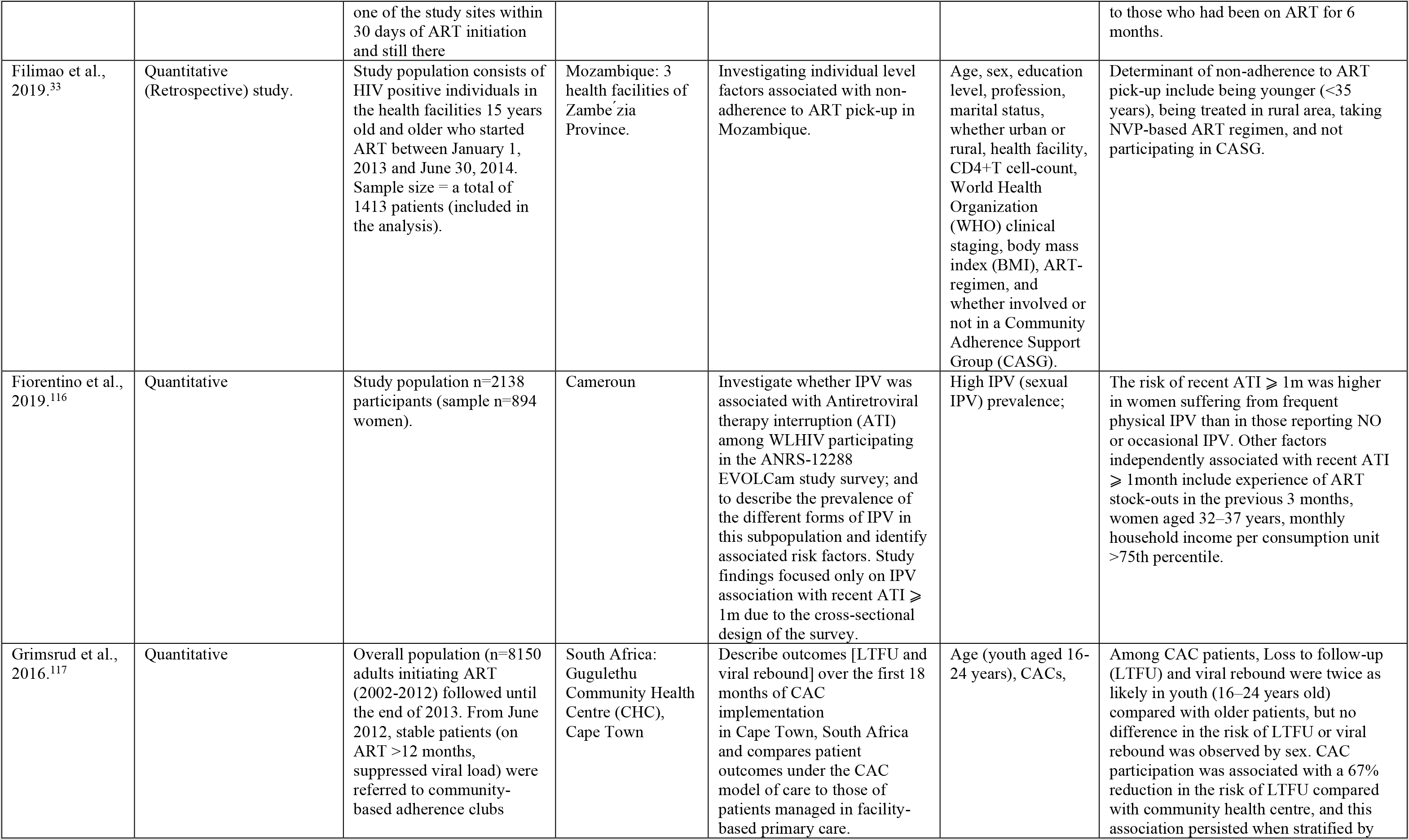
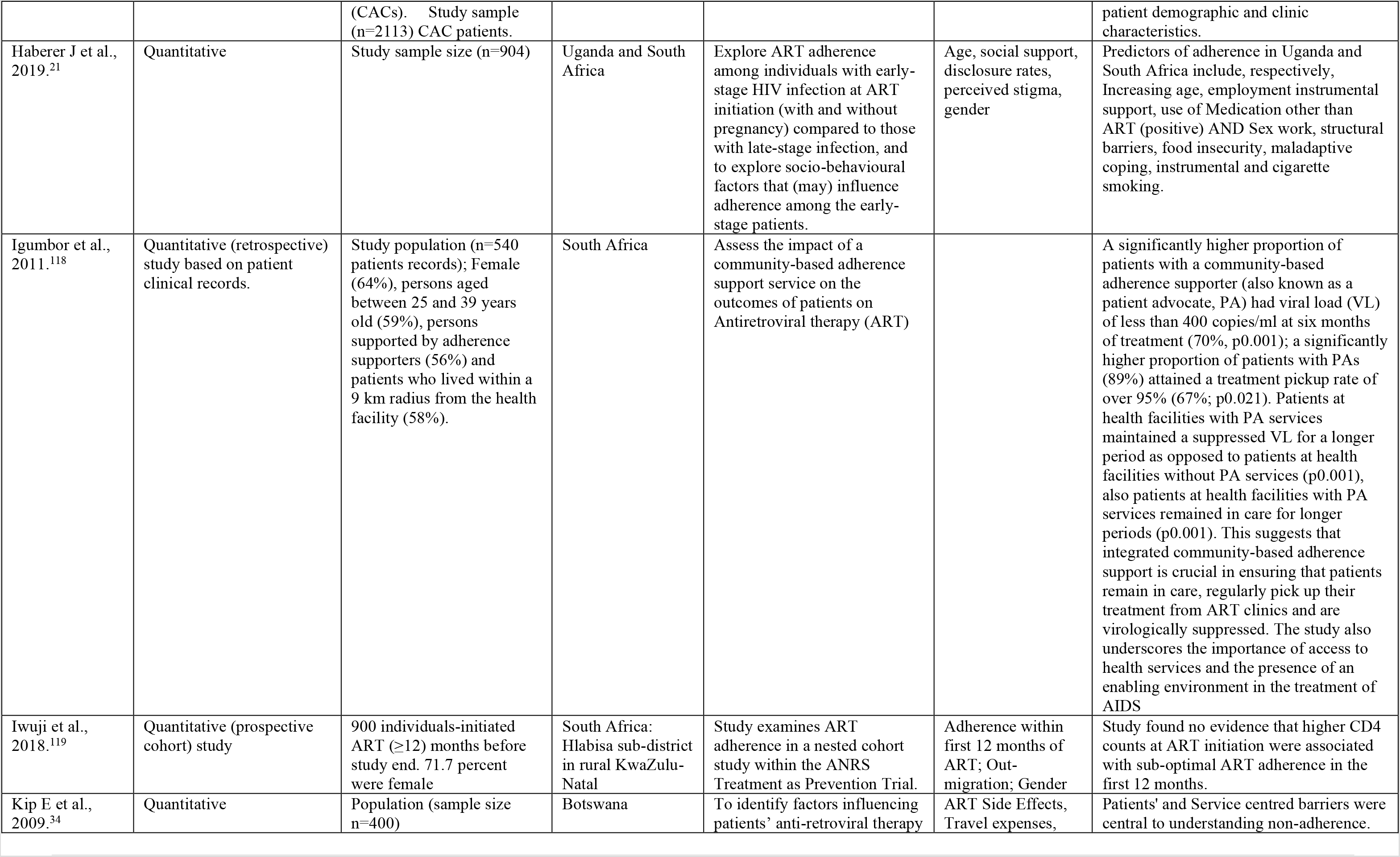
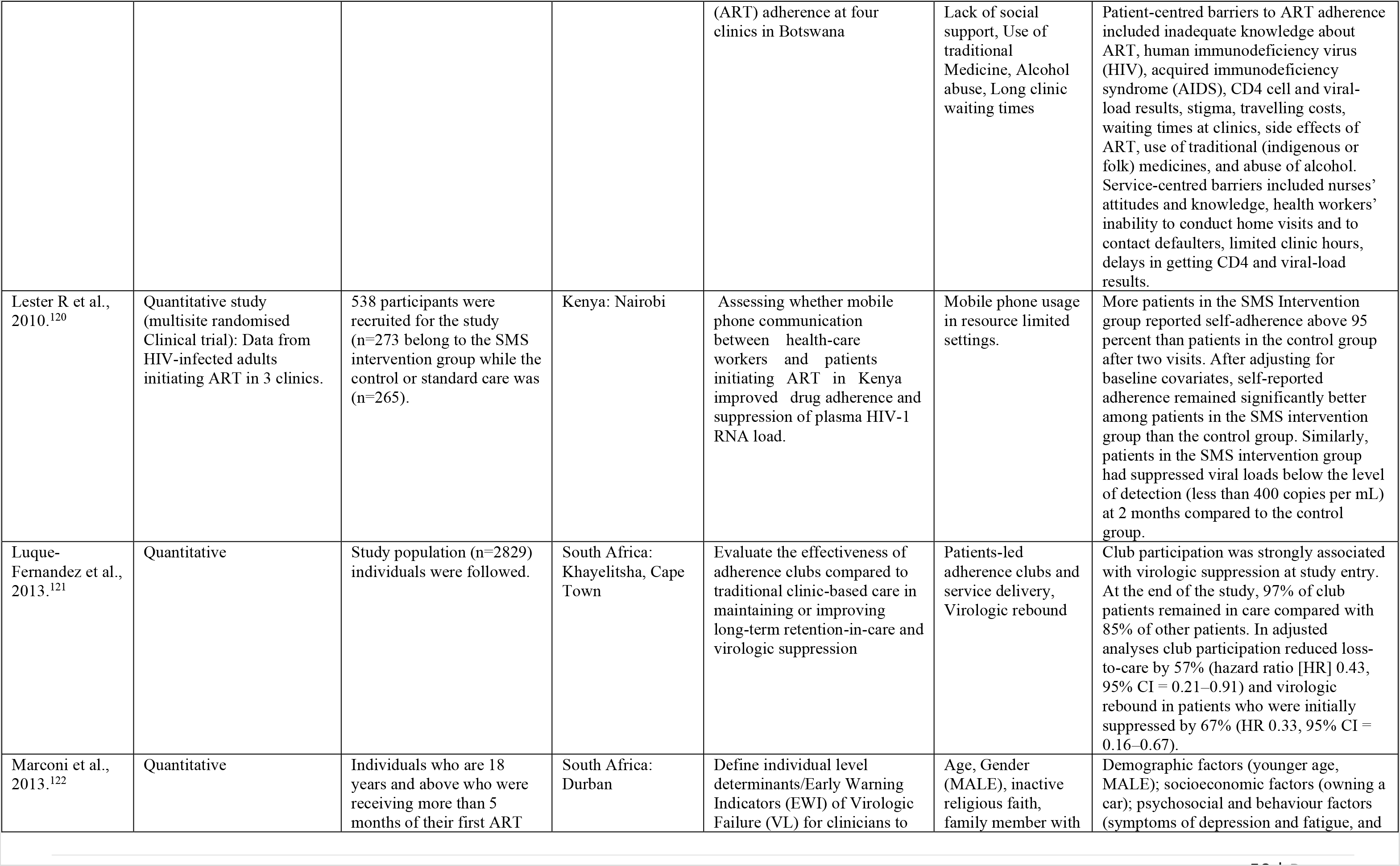
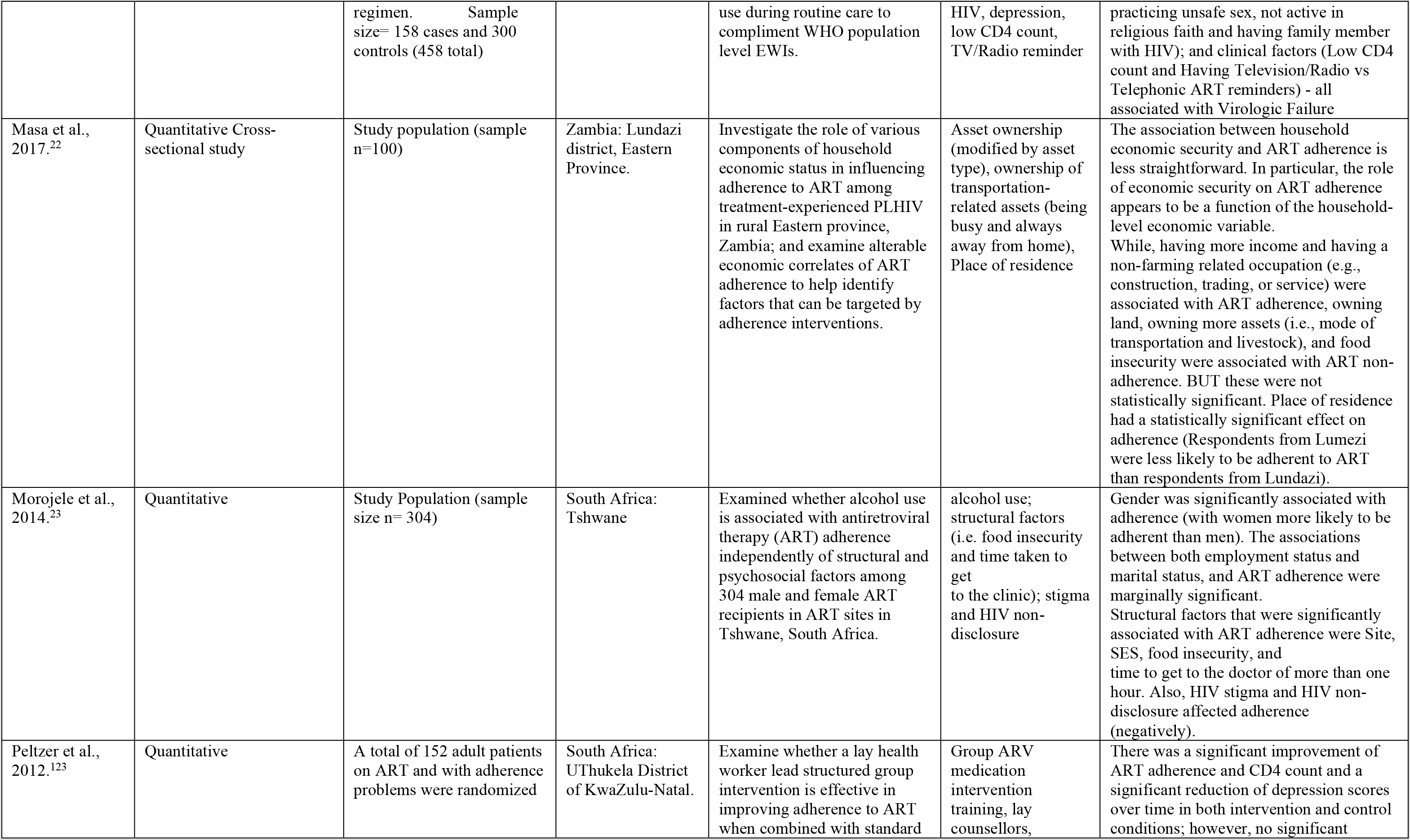
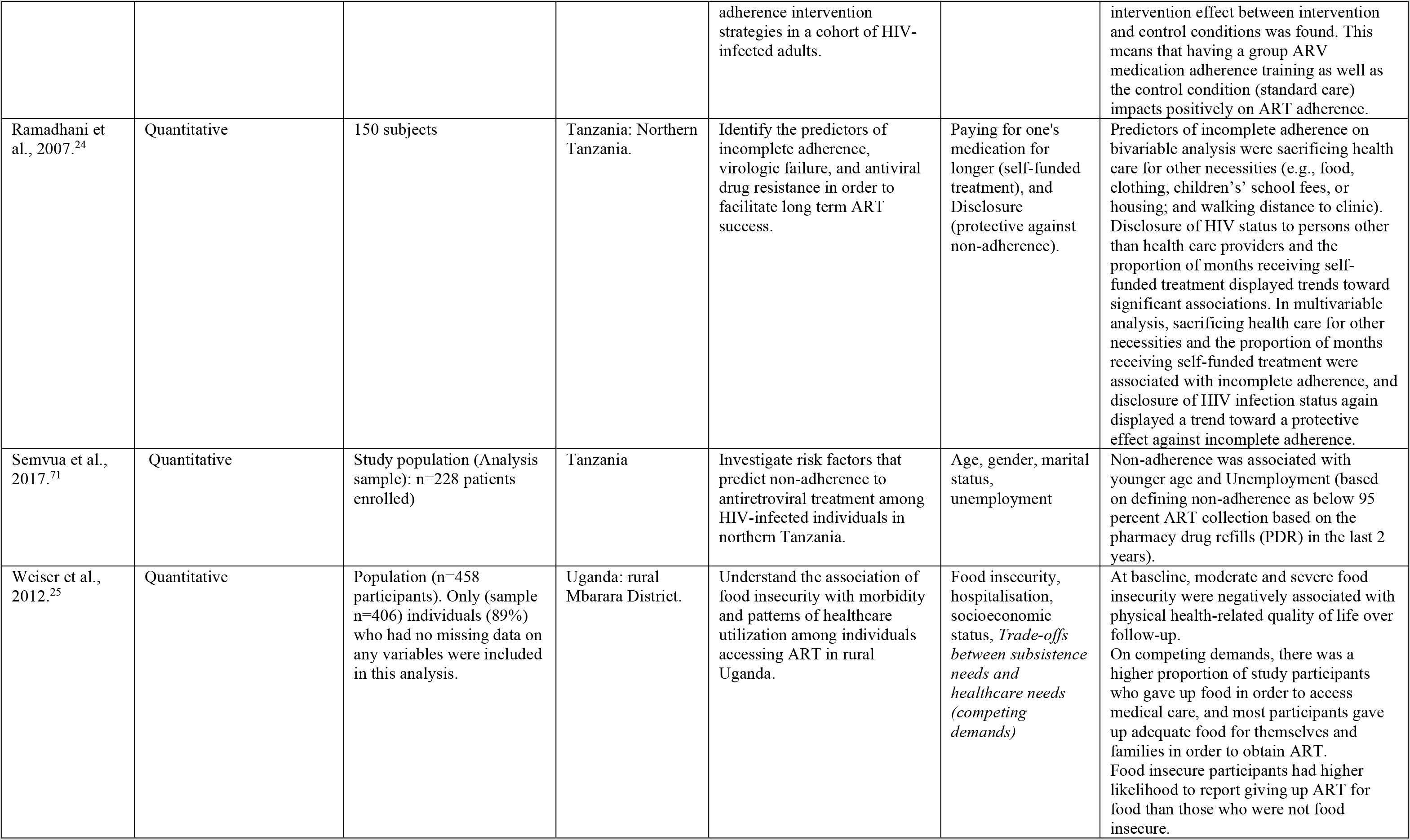
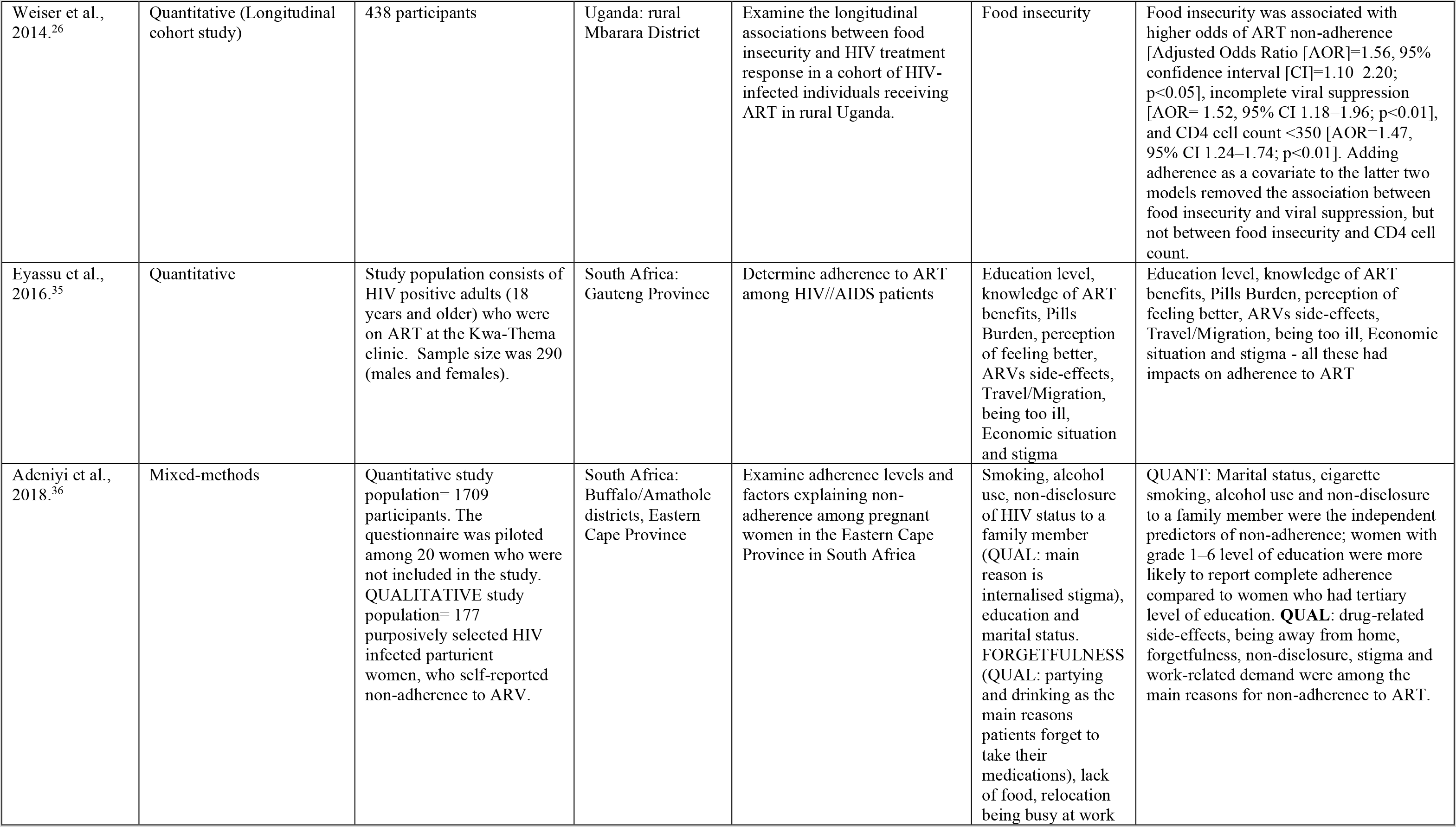
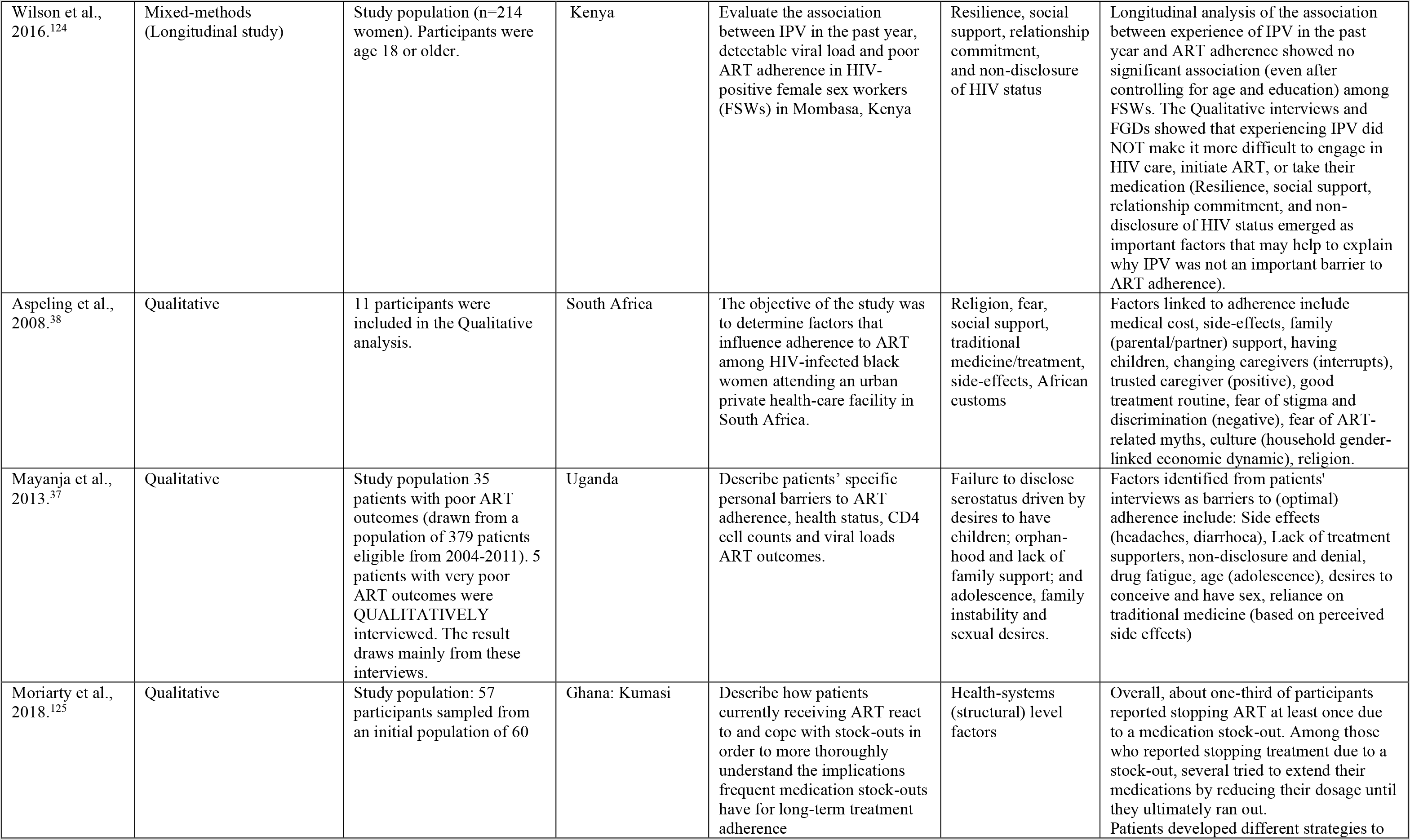
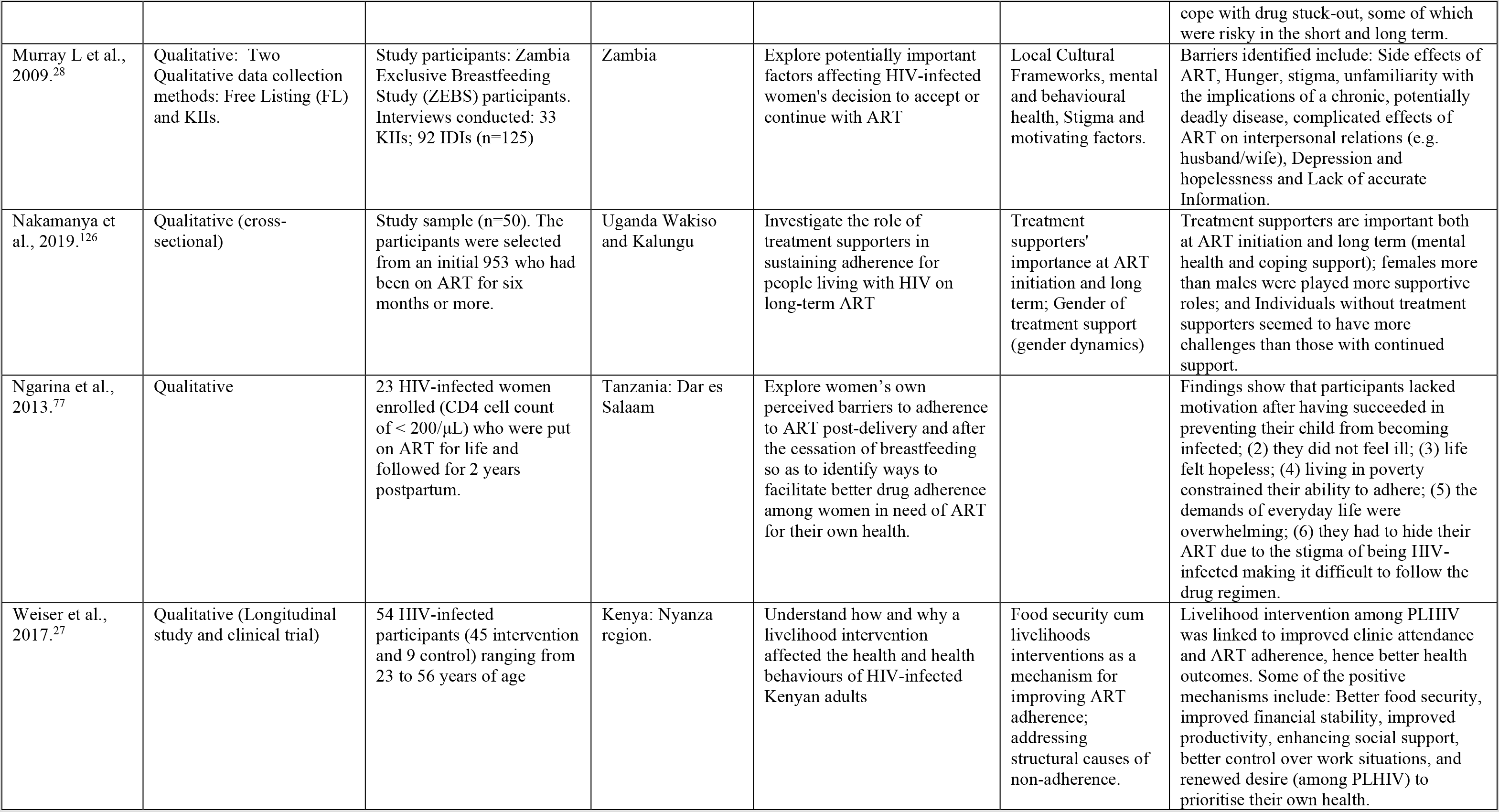
Overview of publications meeting inclusion criteria^5^,^19^,^40^,^41^,^42^,^48^,^49^,^52^,^54^,^56^,^58^,^65^,^67^,^69^,^72^,^73^,^78^,^81^,^82^,^94^,^97^,^120^,^129^,^130^

We used the empirically derived themes (Table 4) from the quantitative and qualitative studies to develop a systems explanation of the relationship between drought and adherence (Figure 2) whilst citing areas of similarity between the theoretical framework such as the socio-ecological model described earlier.

**Figure 2:**
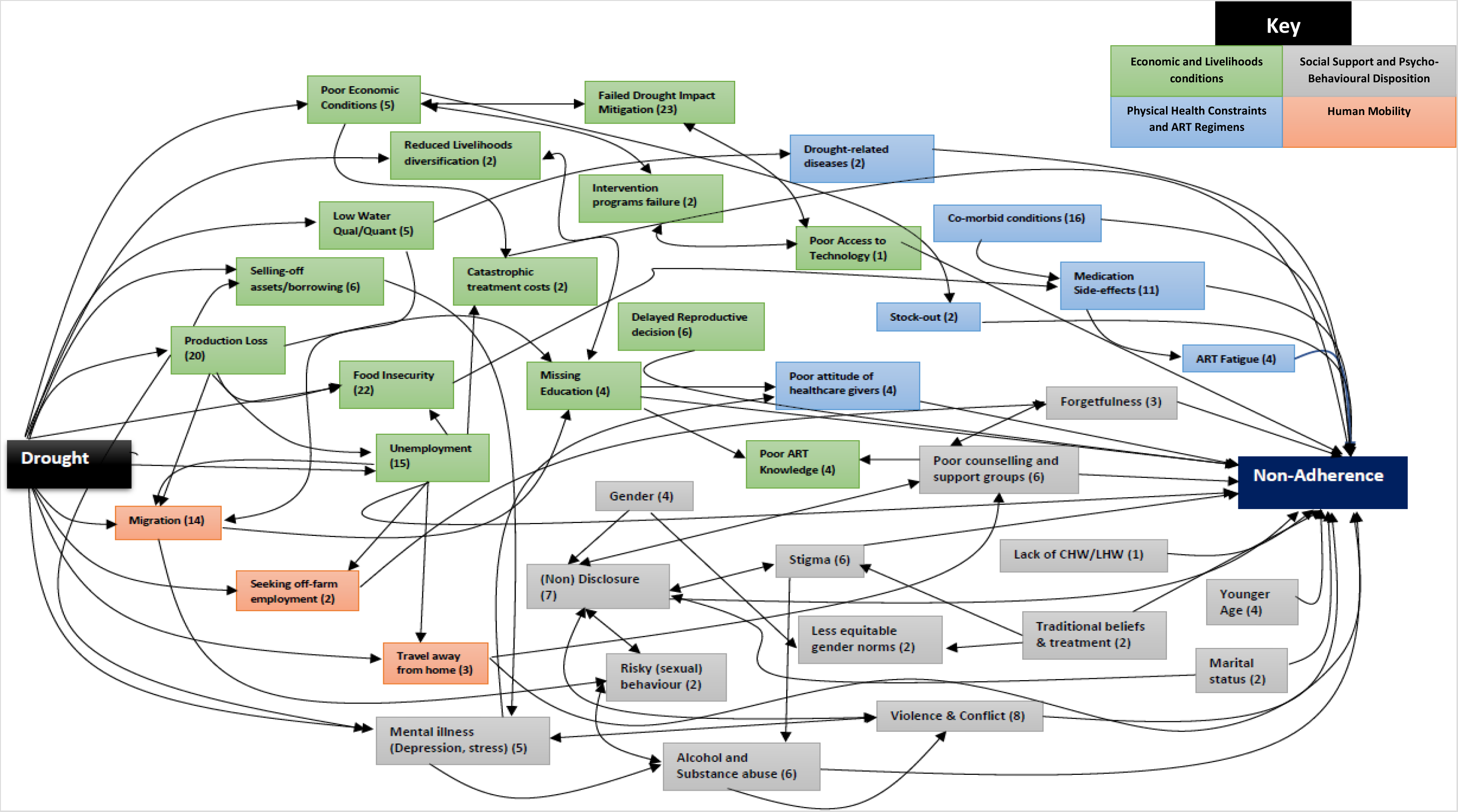
Systems Diagram linking drought and ART non-adherence in Africa. This systems diagram demonstrates the complex interlinkages between drought and ART non-adherence moderated by different factors. The colour codes represent different themes (green = Livelihoods and Economic conditions; grey = social support and psycho-behavioural disposition; light blue = Physical health constraints and ART regimens; orange = human mobility). The numbers represent the number of articles referencing a particular factor in this system

The four thematic areas, a) livelihoods and economic conditions, b) physical health constraints and ART regimens, c) human mobility d) social support and psych-behavioural dispositions (Table 4), illuminate the complex pathways to understanding the extent to which adherence to HIV care is sensitive to the effects of drought on human livelihoods, interactions as well as structures of policy response to environmental stress.

### 3.1 Livelihoods and Economic conditions

Livelihoods and economic conditions emerged as one of strongest determinants of (non)adherence. Factors identified here (Table 4) include food and water insecurity – linked to drought impacts on production, livelihoods and private and public incomes that – cumulatively affected ART adherence. Multiple studies have shown poor socio-economic conditions to be associated with poor adherence. Often cited in the literature is the impact of worries about taking ART on an empty stomach.^21–28^ The fear of adverse side effects linked to food insecurity was also noted as an important barrier to adherence in both quantitative and qualitative studies.^29–38^

Study participants attributed side effects such as hallucinations, drowsiness and sickly feelings to taking ART with insufficient food, or on an empty stomach.^28,36,38^

Drought directly impacts food security through loss of production of both crop and livestock, and for subsistence farmers may affect their capacity to access food.^5,39–70^ Individuals could also be impacted indirectly either through loss of employment, or the increase in food prices.^22,46,71–74^

In response to a severe economic impact of drought, some individuals sold off assets, including those that help to meet individual and family food needs.^51,57,70,75^ In the case of an extended drought, beyond a year or growing season, this may affect mental health (anxiety, stress, depression) which in turn could impact not only individual, but also family socio-economic conditions if the breadwinner in the family is affected.^76^ ART adherence could then be affected through trading off family food provision with the cost of transportation to a healthcare facility for drug pick up or vice versa.^31,36^ However, on the other hand, individuals that are employed and or well-off sometimes find themselves defaulting on treatment due to the demands of work.^22,24,25,31,34,36,77^

Lack of access to clean water, or the means to buy it, is a socioeconomic condition (Table 4) also found to have severe impact on ART adherence.^26,115^ Extreme drought can further exacerbate an already limited resource. Figure 2 shows how low water quality and quantity is linked to diseases afflicting livestock and human beings alike, for example drought-tolerant tuber crops like cassava can lead to Konzo disease.^53,56,59,78,82,102^ (Konzo disease is a type of paralysis of the leg which is permanent and is associated with the consumption of inadequately processed cassava-based food.)^59^ People on ART may also forgo hospital appointments, as they search for clean water for themselves and their livestock, while insufficient and low water quality (driven by drought) may exacerbate poor economic conditions that individuals, communities, farmers, herders and countries face in general.^8,43,46,62,95,99^ Insufficient water impacts overall food production, increases (government) expenditure on food and could exacerbate morbidity. Such fiscal burdens on the economy linked to food and water could have ripple effects on expenditure on adequate water supply to poor drought-stricken (rural) communities where many HIV positive individuals reside. This would inadvertently impact ART adherence in these communities; with poor individuals who rely on government’s provision of water, often inconsistent, being disproportionately affected.

Poor economic conditions, possibly from drought-impact on livelihoods or overarching poverty, impede people’s ability to acquire even the simplest technological device (mobile phones), often used to facilitate ART adherence. Developing technology-based interventions – either for drought-monitoring or ART adherence – heavily depends on economic viability but in many African countries, harsh economic conditions put governments in tough positions to make trade-offs between health interventions and economic stability, amongst others. Consequently, poor economic conditions and insufficient interventions interact negatively to exacerbate how drought affects the agency of individuals and communities.

The economic shocks associated with drought, especially in terms of seasonal poverty, can impact investment in human capital, including education,^45,104,108^ and access to adequate health care. Previous ART adherence studies showed that HIV-positive individuals who had to bear the cost of treatment, including those linked to transportation to clinics, reported poor adherence.^31^ Various support systems,^28,34–36,123^ including community health workers, family members, lay health workers and counselling groups within a drought-hit context may face challenges from drought that impact on their ability to provide support for healthcare in general.

### 3.2 Comorbidities and ART regimen

Drought aggravates the physical and mental health pressures that individuals face.^4–6,8,47,57,59,102,104,105,127^ Like other environmental stressors and extreme events, including floods, drought is linked to disease outbreaks such as Rift Valley Fever, Konzo disease, trachoma, diseases linked to poor hygiene and access to water like diarrhoea, and those carried by vectors like chikungunya outbreaks in East Africa.^4–6,59^ Further, drought has been linked to later life disability, especially among males who also are at risk of both physical and mental disabilities having experienced drought as infants.^105^

It is established that drought imposes economic distress and stress on individuals due to losses in crop and animal production, and livelihoods.^8,57,61^ Such stressful situations, including those linked to income loss, unemployment, seeking off-farm employment or migration, and possibly exacerbated by drought-related diseases and disabilities, may culminate in coping mechanisms that include alcohol and substance abuse which have been implicated in domestic and intimate partner violence (IPV).^13^ Further, migration has been linked to increased risky (sexual) behaviours that culminate in increased prevalence of HIV in Southern Africa.^8^

Alcohol and substance abuse cases have been shown to be associated with comorbidities like diabetes, heart diseases, hepatitis, hypertension and strokes.^128–131^ Treatment for multi-morbidity related to these conditions in addition to acute stress and depression, result in increased pill burden which is associated with poor adherence^6^ and increased likelihood of drug-drug interactions with HIV drugs.^32,34,35,37,38,109,110,119^ This could result in possible trade-off adherence to one treatment over another. In drought, especially among the rural poor, this trade-off may be exacerbated by the lack of sufficient food and clean water further increasing disease susceptibility. Where drought and co-morbidities interact with economic stress, social vulnerabilities, medication stock-out, pills burden or side-effects from ART regimens, this can be detrimental for adherence.^28–38,116,125^

Non-adherence, a negative health seeking behaviour, is also attributable to patients’ experience of, and relationship with their, healthcare providers.^28,30,36,77^ Three studies cited the attitudes of nurses towards patients as a possible barrier to ART adherence, including not trusting health facilities to maintain confidentiality.^23,34,36^ This could be an additional disincentive in patients already facing increased livelihood challenges from drought especially among men who, facing a dominant masculine normativity of breadwinner, feel ashamed, isolated and refuse to seek help for their mental illness due to the stigma attached to mental health.^13^ Drought’s impact on the macro-economy may arguably also exacerbate drug stock-out of more expensive ART regimen with fewer side-effects, as countries may rely on cheaper older regimen with more side effects due to competing policy priorities.

### 3.3 Human mobility

Human mobility, with mobility defined broadly to encapsulate migratory activities as well as other forms of movements, brings together factors that can influence adherence. Many studies showed that adherence is very sensitive to mobility: people moving from where they are resident and registered with healthcare facilities to new and possibly unfamiliar places.^35,114,115,119^ Some included studies showed that drought is a very strong driver of human migration with people moving away from drought affected areas or relocating their livestock to areas where they could find forage and water to prevent them from dying.^7,8,60,84,85,88,90,95,103^ The impact of drought on adherence could be mediated through this forced mobility.

Collectively, these studies show how forced migration from drought or other forms of migration can affect adherence. For example, permanent migration (change of residence) – where people changed locations and lost touch with their primary clinics – or travels outside normal areas of residence for work (including holidays or religious activities of some sort) affected adherence among patients.^114,115^ Similarly, movements generally linked to seeking off farm employment or permanent relocation out of a drought-stricken area were found in relation to the impact of drought on livelihood.^51,64,86,98^

Drought has been linked to violence and displacements as well as increased risky sexual behaviour and alcohol and substance abuse, which have been shown to be associated with poor adherence.^8^ Firstly, the scramble over water sources by herders and farmers is well recorded in countries like Kenya and Nigeria.^86,107^ Although the impact of drought has not been explored in-depth in the Nigerian case, in Kenya, incidents of violence linked to the practice of cattle-rustling emanate from scarce water resources.^80,90,106,107^ In contexts where such violence lead to the displacement of communities, forced migration, the implication for HIV positive individuals in care becomes dire. Secondly, people who migrate in search of better life opportunities out of drought-stricken areas face uncertainties in their destinations that have culminated in many risky sexual behaviours such as transactional sex and alcohol abuse, both of which are strongly associated with poor adherence.^7,8^

Droughts cause crop failures, production losses, livestock deaths or reduced productivity and a (near) total destruction of individual and community livelihoods to the extent that social structures (social networks) within socially-knit communities become stretched.^44^ People move, temporarily, permanently, internally or internationally, to seek out avenues to survive. We found these to include taking refuge outside the drought area, sending children to more affluent relatives (maybe outside the area), sending family members abroad,^44,60,88,96^ selling off their assets to survive, as well as abandoning (rural) farms to seek off-farm employments in cities or even, for cattle farmers, moving their herd away in search of forage in other towns or areas.^44,51,53,64,74,84,85,96^ These outcomes linked to drought, as our review found, are some of drivers of poor adherence.

Consequently, the mitigation strategies established to manage drought impacts were individual-based – moving or sending children or family members to relatives or abroad – and community-focused such as community food or loan support systems, and even on an institutional/policy level of intervention.^44,68,92,95,96,101^ An important issue about the latter is that if, and once, drought succeeds in destabilising support systems set up individually or collectively – within communities and beyond – then the devastation on wealth and health further diminishes community resilience.^44,96^ This is central to the next theme; support systems and the linkages to drought and adherence.

### 3.4 Social support and psycho-behavioural disposition

Our review has shown that migration can mean the loss of (important) support structures, especially strongly knit (community/family) social support systems. This could be a source of anxiety and stress relating to concerns around adjustment and integration in the new environment.^13^ The place and role of support systems for adherence is well-documented in the ART adherence literature. In fact, issues linked to support systems were described in 17 articles highlighting how marital status, non-disclosure and forgetfulness drive poor adherence, and how caregivers’ roles, counselling groups, community or lay health workers (CHW or LHW) were crucial towards facilitating adherence.^23,24,31,33,36–38,109,110,112,113,115,117,118,121,123,124,126^ Conversely, failures in the support system, for example absence of caregivers’ or lack of health workers, were shown detrimental to ART adherence in different population groups, with younger age groups being more affected.^32,33,71,117^

The sensitivity of adherence to the support systems appeared to be exacerbated by gender, which is similarly impacted by drought.^23,45,61,119^ Indeed, where support systems are lacking, livelihood losses induce stress that lead to alcohol and substance abuse and associated risky sexual behaviour, including transactional sex, multiple sexual partners and sex without condom due to weak bargaining power (for women and girls) which studies showed to hinder adherence.^21,32,34,36,122,7,76^ Very importantly, these can emanate from attempts to cope with HIV-related stigma and unpalatable experiences from health workers.^23,28,30,38,77^ Some of these negative coping strategies may inadvertently lead to domestic violence (including IPV), in some cases which also negatively impacts adherence.^116,124^

Social, cultural and religious (gender) norms that normalise such systems of stigmatisation, especially in paternalistic African societies, exacerbate this situation.^23^ Drought, by increasing vulnerability within affected communities, constrains (or possibly erodes) whatever safety nets and support structures that may exist.^44,95,96^ It imposes additional shocks in a situation where poverty already disrupts individuals’ capacity to support themselves or extended family members who may be dependent on them for sustenance.

## 4. Discussion and Conclusion

The individual and public health consequences of poor ART adherence, and resulting increase in HIV drug resistance, in HIV-positive individuals have been clearly described.^132^ At the individual level, these include increased HIV-related morbidity and mortality whilst at the public health level, there is the risk of transmission of, possibly drug resistant, HIV to sexual partners and threat to national HIV treatment programmes based on the public health approach as in many African countries.^133^

In this systematic review, we utilised a systems approach to examine the complex linkages between drought and ART adherence which is mediated through a web of proximate and distal factors not often considered. We found that the strongest links between drought and poor ART adherence were those clustered around livelihoods and economic conditions, with most emphasis on food insecurity, loss of production and individual, community and national economic conditions in the form of unemployment and reduced overall income. These factors interact with social support systems, psycho-behavioural dispositions, mobility, as well as physical health and ART regimen-related constraints in a disruptive manner culminating in varying forms of poor adherence.^13^

Our systems diagram connects these factors, demonstrating adherence sensitivity to drought-related impacts. So, elements of poor adherence, like medication side-effects, comorbidities and migration, are shown to be largely products of constrained livelihood and economic conditions. Such conditions have also been shown to exacerbate stress, depression, stigma, alcohol and substance misuse, risky sexual behaviour, and IPV.^13,134^ Consequently, non-disclosure and or forgetfulness, and inadequate support structures, prove detrimental to adherence.

The reviewed literature described individual, community and institutional strategies to mitigate economic situations and address poor adherence. Institutional and policy frameworks related to drought and environmental stress mitigation include water harvesting, community loan systems, food aid, drought tolerant crops, off-farm employment or alternative livelihoods amongst others.^50,51,53,55,57,63,66,74,75,79,83,86,87,89,91–93,96–98,100,101^ The review also shows the disruptive effects of drought on these strategies.^44,96^

Adherence studies allude to policy and health system factors and their impact on (improving) adherence. Statistical evidence of associations (and good Qualitative narratives from some of these studies) between health systems interventions and positive adherence outcomes among patients have been found in Quantitative studies.^27,123^ This means that changes in medication (ART) regimens, improved policy guidelines for patients’ handling or engagement (targeted at care providers and facilities) as well as sentiments of trust towards care providers, the effectiveness of counselling/support groups and community/lay health workers signalled improvements in adherence outcomes in the reviewed literature. This highlights the essential role of policy and health systems as regards these positive outcomes. Conversely, failure within these systems is detrimental to adherence.

Noteworthy, and linked to drought impact mitigation, is that poor economic conditions – a possible impact of drought – can detract institutional frameworks’ attempts to address the challenges of livelihood loss(es) and adherence. Because drought’s effects strike deep into the economy, it could raise the country’s debts and increase the opportunity costs that may truncate strides to cushion both environmental stress and adherence challenges.^13^ A strong and resilient economy is more favourable towards advancing ART adherence and limiting its negative consequences by ensuring sufficient medication supplies, reviewing and reforming medication regimens with increased adverse effects on patients while also enhancing and supporting food supply systems and various networks of support for HIV positive individuals.

This review, bringing together environmental and physical related factors linked to drought and various barriers and facilitators of ART adherence in Africa, demonstrates the strength of a systems approach. The triangulation of quantitative, qualitative and mixed-methods studies enhanced the ability of this study to elucidate complex connections between drought and adherence that were not immediately apparent. This is crucial for future studies on the interaction between drought and HIV-related treatment and adherence challenges. Our review did not include grey literature and studies not published in English, some of which might have been relevant for this review.

## Gaps in literature: towards further research

One major finding from this systematic review is the lack of studies directly investigating the relationship between drought and ART adherence. The systems approach adopted by our review substantially extends the literature in this field by exploring the non-linear and complex pathways between drought and HIV treatment adherence.

The cross-sectional nature of the two studies that have examined the relationship between drought and HIV prevalence meant that the association could not be considered causal, as acknowledged by the authors.^7,8^ Furthermore, HIV prevalence is a weak outcome variable because it is sensitive to the mortality rate and HIV incidence in the population, which in turn is affected by factors other than ART adherence per se. Longitudinal studies investigating the impact of drought on HIV acquisition, and mediators such as population HIV viral load could therefore address the noted shortcomings.

Further, mental health challenges (acute stress, anxiety, depression), as well as stigma, are crucial to understanding drought-adherence nexus since these challenges may exacerbate economic hardship and vice versa.^28,77,111,122,123^ Surprisingly only one article focussed on the impact of drought on mental health in Africa even though stress, broadly speaking, can provide a convergence point in grasping drought’s impact on ART adherence.^76^ The dearth of information in this area highlights the fact that mental health is under-investigated in the African setting especially in the context of medical pluralism in which mental health conditions are often attributed to spiritual or ancestral issues. This is an area that warrants further investigation.

Studies on appropriate mitigation strategies and economic support systems for less resilient economies with a high burden of HIV and individual poverty with limited ability to cushion the impact of drought are urgently required.

## Data Availability

Not applicable

## Acknowledgements

The authors are grateful to the social science department at the African Health Research Institute (AHRI) for their support in the development of this piece of research.

## Authors’ Contributions

All authors approved the final version of the manuscript

| Contributor Role | Role Definition |  |
| --- | --- | --- |
| Conceptualisation | Ideas; formulation or evolution of overarching research goals and aims. | CI, SAK, DK, KO |
| Data Curation | Management activities to annotate (produce metadata), scrub data and maintain research data (including software code, where it is necessary for interpreting the data itself) for initial use and later reuse. | CI, SAK, DK, KO |
| Formal Analysis | Application of statistical, mathematical, computational, or other formal techniques to analyse or synthesise study data. | CI, SAK, DK, KO |
| Funding Acquisition | Acquisition of the financial support for the project leading to this publication. | CI, DK |
| Investigation | Conducting a research and investigation process, specifically performing the experiments, or data/evidence collection. | CI, SAK, DK, KO |
| Methodology | Development or design of methodology; creation of models. | CI, SAK, DK, KO |

| **Contributor Role** | **Role Definition** |  |
| --- | --- | --- |
| Project Administration | Management and coordination responsibility for the research activity planning and execution. | CI, SAK, DK, KO, SN |
| Resources | Provision of study materials, reagents, materials, patients, laboratory samples, animals, instrumentation, computing resources, or other analysis tools. | CI, KO |
| Supervision | Oversight and leadership responsibility for the research activity planning and execution, including mentorship external to the core team. | CI |
| Validation | Verification, whether as a part of the activity or separate, of the overall replication/reproducibility of results/experiments and other research outputs. | KO, SAK, DK, CI |
| Visualization | Preparation, creation and/or presentation of the published work, specifically visualization/data presentation. | KO, CI, DK, SAK, NN, KB, JS, KH, FT, WH |
| Writing – Original Draft Preparation | Creation and/or presentation of the published work, specifically writing the initial draft (including substantive translation). | KO, SAK, DK, CI |
| Writing – Review & Editing | Preparation, creation and/or presentation of the published work by those from the original research group, specifically critical review, commentary or revision – including pre- or post-publication stages. | KO, SAK, DK, CI, KH, KB, JS, NN, FT, WH |

## Data Availability Statement

Not applicable.

## Reference

1 WHO. COP24 special report: health and climate change. Report No. 978-92-4-151497-2, (World Health Organization, 2018, Geneva, 2019).

2 Watts, N. et al. The Lancet Countdown: tracking progress on health and climate change. Lancet 389, 1151–1164 (2017).

3 Stanke, C., Kerac, M., Prudhomme, C., Medlock, J. & Murray, V. Health effects of drought: a systematic review of the evidence. PLoS currents 5 (2013).

4 Anthonj, C., Diekkruger, B., Borgemeister, C. & Kistemann, T. Health risk perceptions and local knowledge of water-related infectious disease exposure among Kenyan wetland communities. International Journal of Hygiene and Environmental Health 222, 34–48 (2019).

5 Anyamba, A. et al. Recent weather extremes and impacts on agricultural production and vector-borne disease outbreak patterns. PloS one 9, e92538 (2014).

6 Anyamba, A. et al. Climate Teleconnections and Recent Patterns of Human and Animal Disease Outbreaks. Plos Neglected Tropical Diseases 6 (2012).

7 Low, A. J. et al. Association between severe drought and HIV prevention and care behaviors in Lesotho: A population-based survey 2016-2017. PLoS Med 16, e1002727 (2019).

8 Burke, M., Gong, E. & Jones, K. Income Shocks and HIV in Africa. Economic Journal 125, 1157–1189 (2015).

9 Clover, J. Food Security in Sub-Saharan Africa. African Security Review 12, 5–15 (2003).

10 UNECA. Sustainable Development Goals for the Southern Africa subregion: Summary report. 28 (United Nations Economic Commission for Africa, Addis Ababa, Ethiopia, 2015).

11 UNAIDS. Global report: UNAIDS report on the global AIDS epidemic 2010. Report No. 9291738719 (Joint United Nations Programme on HIV/AIDS (UNAIDS), 2010).

12 Berry, H. L., Waite, T. D., Dear, K. B. G., Capon, A. G. & Murray, V. The case for systems thinking about climate change and mental health. Nature Climate Change 8, 282–290 (2018).

13 Vins, H., Bell, J., Saha, S. & Hess, J. J. The Mental Health Outcomes of Drought: A Systematic Review and Causal Process Diagram. Int J Environ Res Public Health 12, 13251–13275 (2015).

14 Peters, D. H. The application of systems thinking in health: why use systems thinking? Health research policy and systems 12, 51 (2014).

15 Uganda Ministry of Health. National Antiretroviral treatment care guidelines for Adults and Children (Kampala, Uganda Ministry of Health in Uganda, 2003).

16 CASP. CASP Cohort Study Checklist (Critical Appraisal Skills Programme (CASP), 2018). Accessed 20-June-2020 at <https://casp-uk.net/wp-content/uploads/2018/03/CASP-Cohort-Study-Checklist-2018_fillable_form.pdf>.

17 Katz, I. T. et al. Impact of HIV-related stigma on treatment adherence: systematic review and meta-synthesis. 16, 18640 (2013).

18 Elder, J. P. et al. A description of the social-ecological framework used in the trial of activity for adolescent girls (TAAG). Health education research 22, 155–165 (2007).

19 Mukumbang, F. C., Mwale, J. C. & van Wyk, B. Conceptualising the Factors Affecting Retention in Care of Patients on Antiretroviral Treatment in Kabwe District, Zambia, Using the Ecological Framework. AIDS research and treatment 2017, 7356362 (2017).

20 Stangl, A. L., Lloyd, J. K., Brady, L. M., Holland, C. E. & Baral, S. A systematic review of interventions to reduce HIV-related stigma and discrimination from 2002 to 2013: how far have we come? 16, 18734 (2013).

21 Haberer, J. E. et al. ART adherence and viral suppression are high among most non-pregnant individuals with early-stage, asymptomatic HIV infection: an observational study from Uganda and South Africa. Journal of the International Aids Society 22 (2019).

22 Masa, R., Chowa, G. & Nyirenda, V. Barriers and facilitators of antiretroviral therapy adherence in rural Eastern province, Zambia: the role of household economic status. Ajar-African Journal of Aids Research 16, 91–99 (2017).

23 Morojele, N. K., Kekwaletswe, C. T. & Nkosi, S. Associations between alcohol use, other psychosocial factors, structural factors and antiretroviral therapy (ART) adherence among South African ART recipients. AIDS and behavior 18, 519–524 (2014).

24 Ramadhani, H. O. et al. Predictors of incomplete adherence, virologic failure, and antiviral drug resistance among HIV-infected adults receiving antiretroviral therapy in Tanzania. Clinical Infectious Diseases 45, 1492–1498 (2007).

25 Weiser, S. D. et al. Food insecurity is associated with morbidity and patterns of healthcare utilization among HIV-infected individuals in a resource-poor setting. Aids 26, 67–75 (2012).

26 Weiser, S. D. et al. Longitudinal assessment of associations between food insecurity, antiretroviral adherence and HIV treatment outcomes in rural Uganda. Aids 28, 115–120 (2014).

27 Weiser, S. D. et al. Changes in Health and Antiretroviral Adherence Among HIV-Infected Adults in Kenya: Qualitative Longitudinal Findings from a Livelihood Intervention. AIDS and behavior 21, 415–427 (2017).

28 Murray, L. K. et al. Barriers to acceptance and adherence of antiretroviral therapy in urban Zambian women: a qualitative study. Aids Care-Psychological and Socio-Medical Aspects of Aids/Hiv 21, 78–86 (2009).

29 Elafros, M. A. et al. Patient-Reported Adverse Effects Associated with Combination Antiretroviral Therapy and Coadministered Enzyme-Inducing Antiepileptic Drugs. The American journal of tropical medicine and hygiene 96, 1505–1511 (2017).

30 Denison, J. A. et al. Incomplete adherence among treatment-experienced adults on antiretroviral therapy in Tanzania, Uganda and Zambia. Aids 29, 361–371 (2015).

31 Ehlers, V. J. & Tshisuyi, E. T. Adherence to antiretroviral treatment by adults in a rural area of Botswana. Curationis 38 (2015).

32 Elul, B. et al. High Levels of Adherence and Viral Suppression in a Nationally Representative Sample of HIV-Infected Adults on Antiretroviral Therapy for 6, 12 and 18 Months in Rwanda. PloS one 8 (2013).

33 Filimao, D. B. C. et al. Individual factors associated with time to non-adherence to ART pickup within HIV care and treatment services in three health facilities of Zambezia Province, Mozambique. PloS one 14 (2019).

34 Kip, E., Ehlers, V. J. & van der Wal, D. M. Patients’ adherence to anti-retroviral therapy in Botswana. Journal of Nursing Scholarship 41, 149–157 (2009).

35 Eyassu, M. A., Mothiba, T. M. & Mbambo-Kekana, N. P. Adherence to antiretroviral therapy among HIV and AIDS patients at the Kwa-Thema clinic in Gauteng Province, South Africa. African Journal of Primary Health Care & Family Medicine 8 (2016).

36 Adeniyi, O. V. et al. Factors affecting adherence to antiretroviral therapy among pregnant women in the Eastern Cape, South Africa. Bmc Infectious Diseases 18 (2018).

37 Mayanja, B. N. et al. Personal barriers to antiretroviral therapy adherence: case studies from a rural Uganda prospective clinical cohort. African Health Sciences 13, 311–319 (2013).

38 Aspeling, H. E. & van Wyk, N. C. Factors associated with adherence to antiretroviral therapy for the treatment of HIV-infected women attending an urban care facility. International Journal of Nursing Practice 14, 3–10 (2008).

39 Lobell, D. B., Banziger, M., Magorokosho, C. & Vivek, B. Nonlinear heat effects on African maize as evidenced by historical yield trials. Nature Climate Change 1, 42–45 (2011).

40 Lunde, T. M. & Lindtjorn, B. Cattle and climate in Africa: How climate variability has influenced national cattle holdings from 1961-2008. PeerJ 1, e55 (2013).

41 Bartzke, G. S. et al. Rainfall trends and variation in the Maasai Mara ecosystem and their implications for animal population and biodiversity dynamics. PloS one 13 (2018).

42 Angassa, A. & Oba, G. Relating long-term rainfall variability to cattle population dynamics in communal rangelands and a government ranch in southern Ethiopia. Agricultural Systems 94, 715–725 (2007).

43 Call, M., Gray, C. & Jagger, P. Smallholder responses to climate anomalies in rural Uganda. World Dev 115, 132–144 (2019).

44 Hassan, A. G., Fullen, M. A. & Oloke, D. Problems of drought and its management in Yobe State, Nigeria. Weather and Climate Extremes 23 (2019).

45 Hyland, M. & Russ, J. Water as destiny - The long-term impacts of drought in sub-Saharan Africa. World Development 115, 30–45 (2019).

46 Kilimani, N., van Heerden, J., Bohlmann, H. & Roos, L. Economy-wide impact of drought induced productivity losses. Disaster Prevention and Management 27, 636–648 (2018).

47 Yiran, G. A. B. & Stringer, L. C. Spatio-temporal analyses of impacts of multiple climatic hazards in a savannah ecosystem of Ghana. Climate Risk Management 14, 11–26 (2016).

48 Mare, F., Bahta, Y. T. & Van Niekerk, W. The impact of drought on commercial livestock farmers in South Africa. Development in Practice 28, 884–898 (2018).

49 Shi, W. J. & Tao, F. L. Vulnerability of African maize yield to climate change and variability during 1961-2010. Food Security 6, 471–481 (2014).

50 Yengoh, G. T. Climate and Food Production: Understanding Vulnerability from Past Trends in Africa’s Sudan-Sahel. Sustainability 5, 52–71 (2013).

51 Clarke, C. L., Shackleton, S. E. & Powell, M. Climate change perceptions, drought responses and views on carbon farming amongst commercial livestock and game farmers in the semiarid Great Fish River Valley, Eastern Cape province, South Africa. African Journal of Range & Forage Science 29, 13–23 (2012).

52 Codjoe, S. N. A. & Owusu, G. Climate change/variability and food systems: evidence from the Afram Plains, Ghana. Regional Environmental Change 11, 753–765 (2011).

53 Cooper, S. J. & Wheeler, T. Rural household vulnerability to climate risk in Uganda. Regional Environmental Change 17, 649–663 (2017).

54 Derbile, E. K., File, D. J. M. & Dongzagla, A. The double tragedy of agriculture vulnerability to climate variability in Africa: How vulnerable is smallholder agriculture to rainfall variability in Ghana? Jamba 8, 249 (2016).

55 Mthembu, N. N. & Zwane, E. M. The adaptive capacity of smallholder mixed-farming systems to the impact of climate change: The case of KwaZulu-Natal in South Africa. Jamba 9, 469 (2017).

56 Terry, A. K. The impact of the 2015-16 El Nino drought on the irrigated home gardens of the Komati downstream development project, Swaziland. South African Geographical Journal 102, 41–58 (2020).

57 Quinn, C. H., Ziervogel, G., Taylor, A., Takama, T. & Thomalla, F. Coping with Multiple Stresses in Rural South Africa. Ecology and Society 16 (2011).

58 Schmidt, M. & Pearson, O. Pastoral livelihoods under pressure: Ecological, political and socioeconomic transitions in Afar (Ethiopia). Journal of Arid Environments 124, 22–30 (2016).

59 Brown, A. L., Cavagnaro, T. R., Gleadow, R. & Miller, R. E. Interactive effects of temperature and drought on cassava growth and toxicity: implications for food security? Global Change Biology 22, 3461–3473 (2016).

60 Nawrotzki, R. J., Schlak, A. M. & Kugler, T. A. Climate, migration, and the local food security context: introducing Terra Populus. Population and Environment 38, 164–184 (2016).

61 Hennink, M. & McFarland, D. A. A delicate web: household changes in health behaviour enabled by microcredit in Burkina Faso. Glob Public Health 8, 144–158 (2013).

62 Brown, C., Meeks, R., Hunu, K. & Yu, W. Hydroclimate risk to economic growth in sub-Saharan Africa. Climatic Change 106, 621–647 (2011).

63 Dile, Y. T. et al. Assessing the implications of water harvesting intensification on upstream-downstream ecosystem services: A case study in the Lake Tana basin. Science of the Total Environment 542, 22–35 (2016).

64 Gao, J. F. & Mills, B. F. Weather Shocks, Coping Strategies, and Consumption Dynamics in Rural Ethiopia. World Development 101, 268–283 (2018).

65 Twongyirwe, R. et al. Perceived effects of drought on household food security in Southwestern Uganda: Coping responses and determinants. Weather and Climate Extremes 24 (2019).

66 Cafer, A. M. Khat: Adaptive Community Resilience Strategy or Short-Sighted Money Maker? Rural Sociology 83, 772–798 (2018).

67 Nnadi, O. I., Liwenga, E. T., Lyimo, J. G. & Madukwe, M. G. Impacts of variability and change in rainfall on gender of farmers in Anambra, Southeast Nigeria. Heliyon 5 (2019).

68 Speranza, C. I., Kiteme, B. & Wiesmann, U. Droughts and famines: The underlying factors and the causal links among agro-pastoral households in semi-arid Makueni district, Kenya. Global Environmental Change-Human and Policy Dimensions 18, 220–233 (2008).

69 Gwatirisa, P. & Manderson, L. Food Insecurity and HIV/AIDS in Low-income Households in Urban Zimbabwe. Human Organization 68, 103–112 (2009).

70 Lawson, D. & Kasirye, I. How the extreme poor cope with crises: Understanding the role of assets and consumption. Journal of International Development 25, 1129–1143 (2013).

71 Semvua, S. K. et al. Predictors of non-adherence to antiretroviral therapy among HIV infected patients in northern Tanzania. PloS one 12, e0189460 (2017).

72 Silva, J. A. & Matyas, C. J. Relating Rainfall Patterns to Agricultural Income: Implications for Rural Development in Mozambique. Weather Climate and Society 6, 218–237 (2014).

73 Hlahla, S. & Hill, T. R. Responses to Climate Variability in Urban Poor Communities in Pietermaritzburg, KwaZulu-Natal, South Africa. Sage Open 8 (2018).

74 Speranza, C. I. Drought Coping and Adaptation Strategies: Understanding Adaptations to Climate Change in Agro-pastoral Livestock Production in Makueni District, Kenya. European Journal of Development Research 22, 623–642 (2010).

75 Coppock, D. L. & Desta, S. Collective Action, Innovation, and Wealth Generation Among Settled Pastoral Women in Northern Kenya. Rangeland Ecology & Management 66, 95–105 (2013).

76 Bahta, Y. T., Jordaan, A. & Muyambo, F. Communal farmers’ perception of drought in South Africa: Policy implication for drought risk reduction. International Journal of Disaster Risk Reduction 20, 39–50 (2016).

77 Ngarina, M., Popenoe, R., Kilewo, C., Biberfeld, G. & Ekstrom, A. M. Reasons for poor adherence to antiretroviral therapy postnatally in HIV-1 infected women treated for their own health: experiences from the Mitra Plus study in Tanzania. Bmc Public Health 13 (2013).

78 Mussa, F. E. F., Zhou, Y., Maskey, S., Masih, I. & Uhlenbrook, S. Groundwater as an emergency source for drought mitigation in the Crocodile River catchment, South Africa. Hydrology and Earth System Sciences 19, 1093–1106 (2015).

79 Nalley, L., Dixon, B., Chaminuka, P., Naledzani, Z. & Coale, M. J. The role of public wheat breeding in reducing food insecurity in South Africa. PloS one 13 (2018).

80 Jones, B. T., Mattiacci, E. & Braumoeller, B. F. Food scarcity and state vulnerability: Unpacking the link between climate variability and violent unrest. Journal of Peace Research 54, 335–350 (2017).

81 Davies, S. Do shocks have a persistent impact on consumption? The case of rural Malawi. Progress in Development Studies 10, 75–79 (2010).

82 Ferrer, N. et al. Groundwater hydrodynamics of an Eastern Africa coastal aquifer, including La Nina 2016-17 drought. Science of the Total Environment 661, 575–597 (2019).

83 Fisher, M. et al. Drought tolerant maize for farmer adaptation to drought in sub-Saharan Africa: Determinants of adoption in eastern and southern Africa. Climatic Change 133, 283–299 (2015).

84 Gray, C. & Mueller, V. Drought and Population Mobility in Rural Ethiopia. World Development 40, 134–145 (2012).

85 Henry, S., Schoumaker, B. & Beauchemin, C. The impact of rainfall on the first out-migration: A multi-level event-history analysis in Burkina Faso. Population and Environment 25, 423–460 (2004).

86 Linke, A. M., Witmer, F. D. W., O’Loughlin, J., McCabe, J. T. & Tir, J. Drought, Local Institutional Contexts, and Support for Violence in Kenya. Journal of Conflict Resolution 62, 1544–1578 (2018).

87 Makate, C., Makate, M., Mango, N. & Siziba, S. Increasing resilience of smallholder farmers to climate change through multiple adoption of proven climate-smart agriculture innovations. Lessons from Southern Africa. Journal of Environmental Management 231, 858–868 (2019).

88 Nawrotzki, R. J. & Bakhtsiyarava, M. International Climate Migration: Evidence for the Climate Inhibitor Mechanism and the Agricultural Pathway. Population Space and Place 23 (2017).

89 Nonvide, G. M. A., Sarpong, D. B., Kwadzo, G. T. M., Anim-Somuah, H. & Gero, F. A. Farmers’ perceptions of irrigation and constraints on rice production in Benin: a stakeholder-consultation approach. International Journal of Water Resources Development 34, 1001–1021 (2018).

90 Owain, E. L. & Maslin, M. A. Assessing the relative contribution of economic, political and environmental factors on past conflict and the displacement of people in East Africa. Palgrave Communications 4 (2018).

91 Adgo, E., Teshome, A. & Mati, B. Impacts of long-term soil and water conservation on agricultural productivity: The case of Anjenie watershed, Ethiopia. Agricultural Water Management 117, 55–61 (2013).

92 Antwi-Agyei, P., Stringer, L. C. & Dougill, A. J. Livelihood adaptations to climate variability: insights from farming households in Ghana. Regional Environmental Change 14, 1615–1626 (2014).

93 Asare-Kyei, D., Renaud, F. G., Kloos, J., Walz, Y. & Rhyner, J. Development and validation of risk profiles of West African rural communities facing multiple natural hazards. PloS one 12, e0171921 (2017).

94 Baudoin, M. A., Vogel, C., Nortje, K. & Naik, M. Living with drought in South Africa: lessons learnt from the recent El Nino drought period. International Journal of Disaster Risk Reduction 23, 128–137 (2017).

95 Bola, G. et al. Coping with droughts and floods: A Case study of Kanyemba, Mbire District, Zimbabwe. Physics and Chemistry of the Earth 67-69, 180–186 (2014).

96 Bosongo, G. B., Longo, J. N., Goldin, J. & Muamba, V. L. Socioeconomic impacts of floods and droughts in the middle Zambezi river basin Case of Kanyemba. International Journal of Climate Change Strategies and Management 6, 131–144 (2014).

97 Fisher, M. & Carr, E. R. The influence of gendered roles and responsibilities on the adoption of technologies that mitigate drought risk: The case of drought-tolerant maize seed in eastern Uganda. Global Environmental Change-Human and Policy Dimensions 35, 82–92 (2015).

98 Little, P. D. Food aid dependency in northeastern Ethiopia: Myth or reality? World Development 36, 860–874 (2008).

99 Markantonis, V. et al. Assessing floods and droughts in the Mekrou River basin (West Africa): a combined household survey and climatic trends analysis approach. Natural Hazards and Earth System Sciences 18, 1279–1296 (2018).

100 Mavhura, E., Manatsa, D. & Mushore, T. Adaptation to drought in arid and semi-arid environments: Case of the Zambezi Valley, Zimbabwe. Jamba 7, 144 (2015).

101 Unks, R. R., King, E. G., Nelson, D. R., Wachira, N. P. & German, L. A. Constraints, multiple stressors, and stratified adaptation: Pastoralist livelihood vulnerability in a semi-arid wildlife conservation context in Central Kenya. Global Environmental Change-Human and Policy Dimensions 54, 124–134 (2019).

102 Hove, J. et al. ‘Water is life’: developing community participation for clean water in rural South Africa. Bmj Global Health 4 (2019).

103 Simatele, D. & Simatele, M. Migration as an adaptive strategy to climate variability: a study of the Tonga-speaking people of Southern Zambia. Disasters 39, 762–781 (2015).

104 Abiona, O. Adverse Effects of Early Life Extreme Precipitation Shocks on Short-term Health and Adulthood Welfare Outcomes. Review of Development Economics 21, 1229–1254 (2017).

105 Dinkelman, T. Long-run Health Repercussions of Drought Shocks: Evidence from South African Homelands. Economic Journal 127, 1906–1939 (2017).

106 Eissler, S., Thiede, B. C. & Strube, J. Climatic variability and changing reproductive goals in Sub-Saharan Africa. Global Environmental Change-Human and Policy Dimensions 57, 11 (2019).

107 Linke, A. M., O’Loughlin, J., McCabe, J. T., Tir, J. & Witmer, F. D. W. Rainfall variability and violence in rural Kenya: Investigating the effects of drought and the role of local institutions with survey data. Global Environmental Change-Human and Policy Dimensions 34, 35–47 (2015).

108 Randell, H. & Gray, C. Climate variability and educational attainment: Evidence from rural Ethiopia. Global Environmental Change-Human and Policy Dimensions 41, 111–123 (2016).

109 Molemans, M. et al. Changes in disclosure, adherence and healthcare interactions after the introduction of immediate ART initiation: an analysis of patient experiences in Swaziland. Tropical medicine & international health: TM & IH 24, 563–570 (2019).

110 Aboubacrine, S. A. et al. Inadequate adherence to antiretroviral treatment and prevention in hospital and community sites in Burkina Faso and Mali: a study by the ATARAO group. International Journal of Std & Aids 18, 741–747 (2007).

111 Adejumo, O. et al. Psychiatric disorders and adherence to antiretroviral therapy among a population of HIV-infected adults in Nigeria. International journal of STD & AIDS 27, 938–949 (2016).

112 Avong, Y. K. et al. Adherence to Anti-Retroviral Therapy in North Central Nigeria. Current HIV research 13, 268–278 (2015).

113 Brittain, K. et al. Determinants of suboptimal adherence and elevated HIV viral load in pregnant women already on antiretroviral therapy when entering antenatal care in Cape Town, South Africa. AIDS Care, (2018).

114 Carlucci, J. G. et al. Predictors of adherence to antiretroviral therapy in rural Zambia. Jaids-Journal of Acquired Immune Deficiency Syndromes 47, 615–622 (2008).

115 El-Khatib, Z. et al. Adherence and virologic suppression during the first 24 weeks on antiretroviral therapy among women in Johannesburg, South Africa - a prospective cohort study. Bmc Public Health 11 (2011).

116 Fiorentino, M. et al. Intimate partner violence against HIV-positive Cameroonian women: Prevalence, associated factors and relationship with antiretroviral therapy discontinuity-results from the ANRS-12288 EVOLCam survey. Womens Health 15 (2019).

117 Grimsrud, A., Lesosky, M., Kalombo, C., Bekker, L. G. & Myer, L. Implementation and Operational Research: Community-Based Adherence Clubs for the Management of Stable Antiretroviral Therapy Patients in Cape Town, South Africa: A Cohort Study. Journal of acquired immune deficiency syndromes (1999) 71, e16–23 (2016).

118 Igumbor, J. O., Scheepers, E., Ebrahim, R., Jason, A. & Grimwood, A. An evaluation of the impact of a community-based adherence support programme on ART outcomes in selected government HIV treatment sites in South Africa. AIDS Care 23, 231–236 (2011).

119 Iwuji, C. et al. Universal test and treat is not associated with sub-optimal antiretroviral therapy adherence in rural South Africa: the ANRS 12249 TasP trial. Journal of the International AIDS Society 21, e25112 (2018).

120 Lester, R. T. et al. Effects of a mobile phone short message service on antiretroviral treatment adherence in Kenya (WelTel Kenya1): a randomised trial. Lancet 376, 1838–1845 (2010).

121 Luque-Fernandez, M. A. et al. Effectiveness of patient adherence groups as a model of care for stable patients on antiretroviral therapy in Khayelitsha, Cape Town, South Africa. PloS one 8, e56088 (2013).

122 Marconi, V. C. et al. Early warning indicators for first-line virologic failure independent of adherence measures in a South African urban clinic. AIDS patient care and STDs 27, 657–668 (2013).

123 Peltzer, K. et al. Efficacy of a lay health worker led group antiretroviral medication adherence training among non-adherent HIV-positive patients in KwaZulu-Natal, South Africa: Results from a randomized trial. Sahara J-Journal of Social Aspects of Hiv-Aids 9, 218–226 (2012).

124 Wilson, K. S. et al. A Prospective Study of Intimate Partner Violence as a Risk Factor for Detectable Plasma Viral Load in HIV-Positive Women Engaged in Transactional Sex in Mombasa, Kenya. AIDS and behavior 20, 2065–2077 (2016).

125 Moriarty, K., Genberg, B., Norman, B. & Reece, R. The Effect of Antiretroviral Stock-Outs on Medication Adherence Among Patients Living With HIV in Ghana: A Qualitative Study. The Journal of the Association of Nurses in AIDS Care: JANAC 29, 231–240 (2018).

126 Nakamanya, S., Mayanja, B. N., Muhumuza, R., Bukenya, D. & Seeley, J. Are treatment supporters relevant in long-term Antiretroviral Therapy (ART) adherence? Experiences from a long-term ART cohort in Uganda. Global Public Health 14, 469–480 (2019).

127 Bakshi, B., Nawrotzki, R. J., Donato, J. R. & Lelis, L. S. Exploring the link between climate variability and mortality in Sub-Saharan Africa. International Journal of Environment and Sustainable Development 18, 206–237 (2019).

128 Freiberg, M. S. & Kraemer, K. L. Focus on the heart: alcohol consumption, HIV infection, and cardiovascular disease. Alcohol research & health: the journal of the National Institute on Alcohol Abuse and Alcoholism 33, 237–246 (2010).

129 Garfield, L. D. et al. Association of anxiety disorders and depression with incident heart failure. Psychosomatic medicine 76, 128–136 (2014).

130 Shield, K. D., Parry, C. & Rehm, J. Chronic diseases and conditions related to alcohol use. Alcohol research: current reviews 35, 155–173 (2013).

131 Wu, L. T., Zhu, H. & Ghitza, U. E. Multicomorbidity of chronic diseases and substance use disorders and their association with hospitalization: Results from electronic health records data. Drug and alcohol dependence 192, 316–323 (2018).

132 Nachega, J. B. et al. HIV treatment adherence, drug resistance, virologic failure: evolving concepts. Infectious disorders drug targets 11, 167–174 (2011).

133 WHO. Global action plan on HIV drug resistance 2017-2021: 2018 progress report, July 2018: executive summary. (World Health Organization, 2018).

134 Hodgson, I. et al. A systematic review of individual and contextual factors affecting ART initiation, adherence, and retention for HIV-infected pregnant and postpartum women. PloS one 9, e111421 (2014).

